# Wearable sleep staging performance declines with sleep apnea severity and is sensitive to training-set composition in very severe sleep apnea

**DOI:** 10.64898/2026.04.09.26350266

**Authors:** Sho Ogaki, Michiru Kaneda, Tomoyuki Nohara, Syuhei Fujita, Naoshi Osako, Tomoko Yagi, Yasuhiro Tomita, Takanori Ogata

## Abstract

We aimed to evaluate wearable sleep staging across sleep apnea severity, including very severe sleep apnea defined as an apnea–hypopnea index (AHI) ≥ 50 events/h, and to assess how training-set composition affects performance in this subgroup. We analyzed 552 overnight recordings: 318 from the Sleep Lab Dataset and 234 from the Hospital Dataset, of which 26.5% (N=62) had very severe sleep apnea. A deep learning model performed sleep staging from photoplethysmography-derived RR intervals and accelerometry recorded by a wrist-worn device. Baseline performance was assessed by 4-fold cross-validation using randomly partitioned folds from the combined datasets. We examined night-level associations with AHI severity. We also compared the baseline model with an ablation model trained on the same number of recordings but with all Sleep Lab Dataset recordings and lower-AHI Hospital Dataset recordings, evaluating both in the very severe subgroup. For 5-stage classification, Cohen’s kappa was 0.586 in the Sleep Lab Dataset and 0.446 in the Hospital Dataset. Under 4-stage staging, the gap narrowed, with kappa values of 0.632 and 0.525, respectively. In the Hospital Dataset, kappa declined with AHI severity, with median kappa differing by about 0.2 between mild and very severe groups. In the very severe subgroup, kappa decreased from 0.365 (baseline) to 0.303 (ablation). Wearable sleep staging performance tended to decline across greater sleep apnea severity. Clinical utility may benefit from training data spanning the target severity spectrum and staging granularity matched to the intended use.

## 1 Introduction

Sleep plays a critical role in maintaining physical health—including weight management, immune function, and glucose regulation—and in supporting cognitive and mental functions such as learning, memory, and mood regulation [1–4]. Sleep apnea is characterized by recurrent disordered breathing during sleep, leading to intermittent hypoxemia and sleep fragmentation, and is estimated to affect nearly one billion people worldwide [5]. If left untreated, sleep apnea is associated with serious long-term risks, including cardiovascular disease, stroke, metabolic disorders, and reduced quality of life [6, 7]. Sleep apnea severity is commonly indexed by the apnea–hypopnea index (AHI), defined as the number of apnea and hypopnea events per hour of sleep. AHI spans a wide clinical range, from mild cases (5≤AHI<15) to very severe cases exceeding 100 events/h [8]. These long-term risks tend to increase with higher AHI [9–11].

Overnight polysomnography (PSG) is the gold standard for evaluating sleep and diagnosing sleep apnea. However, it is costly, requires trained staff and dedicated equipment, and may disturb natural sleep because multiple sensors are attached, which can induce the so-called first-night effect [12, 13]. In contrast, low-cost and noninvasive wearable devices enable long-term sleep monitoring in home environments and have emerged as promising tools for scalable sleep assessment. In particular, wrist-worn and ring-type wearables equipped with photoplethysmography (PPG) sensors and accelerometers constitute an important category for practical, everyday monitoring [14, 15]. This capability may be particularly valuable for individuals with severe sleep apnea, who face higher long-term cardiovascular and metabolic risks. Repeated home-based assessments may complement single-night PSG evaluations, which can be influenced by night-to-night variability and first-night effects.

With recent advances in machine learning, particularly deep learning, wearable-based sleep staging using PPG and accelerometry has been reported to achieve performance approaching that of PSG-based manual scoring [16–21]. However, many existing models have been developed and validated mostly in healthy or general populations, and relatively fewer studies have focused on individuals with sleep apnea [22, 23]. For example, a review of 35 wearable sleep staging studies reported that only 11% validated performance in both healthy individuals and individuals with sleep disorders, including sleep apnea [23]. Although some prior studies have included participants with sleep apnea, evidence remains limited specifically for very severe sleep apnea with AHI exceeding 50–60. In this subgroup, recurrent respiratory events can trigger frequent arousals and substantial disruption of sleep architecture, resulting in sleep patterns that can differ markedly from those in less severe or non-clinical sleep. Moreover, studies often report only a binary comparison between healthy individuals and individuals with sleep apnea, or they pool all individuals with AHI ≥ 30 into a single “severe” group. Such reporting practices can obscure model robustness in the most clinically burdened subgroup.

From a model-development perspective, it also remains unclear how inclusion of very severe cases in the training data affects performance in this subgroup. In particular, algorithms trained mainly on non-clinical or lower-severity recordings may not adequately capture the autonomic and movement patterns observed in nights with very high respiratory-event burden and fragmented sleep. Taken together, current evidence leaves two main practical gaps for clinical use: limited validation across the full sleep apnea severity spectrum, especially in very severe cases; and limited evidence on how inclusion of very severe cases in the training data affects performance in this subgroup.

In this study, we developed a deep learning approach for 5-stage wearable sleep staging and evaluated it across the sleep apnea severity spectrum, with particular attention to very severe sleep apnea, using a sleep-laboratory dataset and a clinical sleep apnea dataset. The sleep-laboratory dataset served as a non-diagnostic, reference-like cohort, whereas the clinical dataset provided substantial representation of high-severity cases. We used our proprietary wrist-worn device equipped with a PPG sensor and an accelerometer. In this paper, we defined very severe sleep apnea as AHI ≥ 50 events/h. Using this framework, we addressed these gaps with a focus on severity-aware evaluation and target-population-aware model development. Specifically, we aimed to:

1. characterize how night-level wearable sleep staging performance varied across AHI severity, including very severe sleep apnea; and
2. assess how inclusion of very severe cases in the training data affected performance in very severe sleep apnea and derive practical implications for model development for this target population.

## 2 Methods

### 2.1 Wearable Device

We used a custom in-house wrist-worn device equipped with a reflective PPG sensor and a 3-axis accelerometer. RR intervals (RRI) were derived from the reflective PPG signal using an algorithm embedded in firmware running on a dedicated sensor-hub IC. The firmware output RRI values on a 25-Hz timestamp grid at time points where a heartbeat peak was detected, rather than as a uniformly sampled time series. Acceleration was sampled at 25 Hz in the earlier period and at 50 Hz after a firmware update. The wrist-worn device operated continuously while detached from the charging dock. A representative view of the wearable device is provided in Fig. 1.

**Figure 1:**
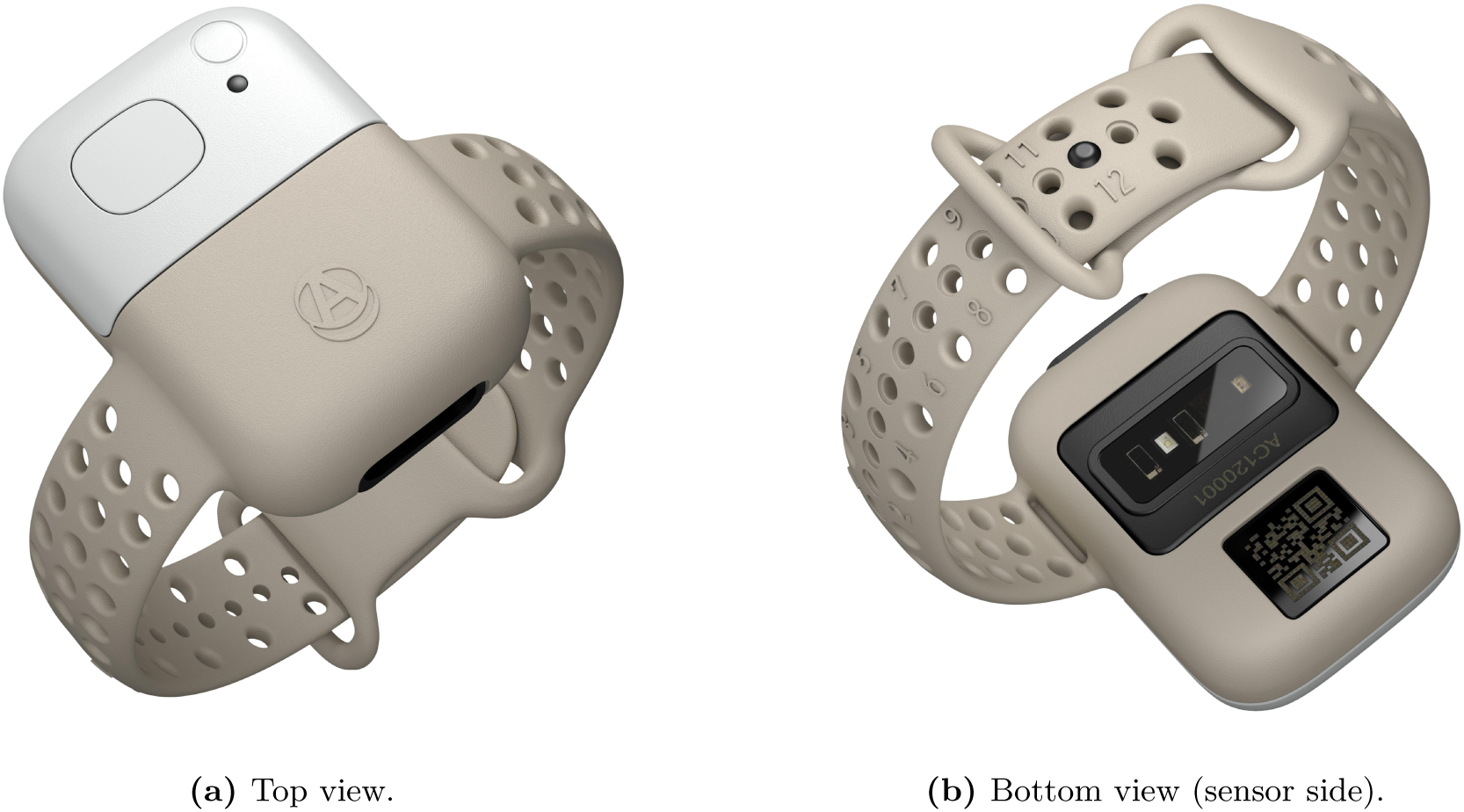
Representative views of the custom in-house wrist-worn device used in this study. Panel (a) shows the top view and panel (b) shows the bottom view (sensor side). The wrist-worn device includes a reflective PPG sensor and a three-axis accelerometer.

### 2.2 Study Design and Datasets

This study analyzed two datasets with simultaneous overnight PSG and wearable recordings. Sleep stages and events were manually scored from the PSG recordings by certified technicians, providing PSG-based reference labels according to the AASM manual [24]. We used a total of 552 overnight recordings from the following two datasets. Participant characteristics and sleep parameters for both datasets are summarized in Table 1.

**Table 1:** Participant characteristics and sleep parameters in the Sleep Lab Dataset and Hospital Dataset.

| Characteristic | Sleep Lab Dataset | Hospital Dataset |
| --- | --- | --- |
| Participant profile |  |  |
| Number of recordings | 318 | 234 |
| Age (years) | 29.7 (11.1) [18, 59] | 56.9 (12.0) [24, 85] |
| Female (N, %) | 149 (46.9) | 48 (20.5) |
| BMI (kg/m<sup>2</sup>) | 21.4 (3.2) [15.6, 37.4] | 25.4 (4.2) [16.1, 45.6] |
| Sleep parameters |  |  |
| TIB (min) | 421.5 (82.5) [133.0, 680.0] | 447.6 (32.9) [346.0, 605.0] |
| TST (min) | 373.9 (87.8) [88.0, 565.0] | 360.4 (56.5) [172.5, 465.0] |
| SE (%) | 88.7 (12.5) [27.6, 99.0] | 80.7 (12.5) [43.4, 97.8] |
| WASO (min) | 39.5 (50.1) [1.5, 259.5] | 71.5 (53.5) [5.5, 266.5] |
| SL (min) | 5.7 (10.7) [0.0, 118.0] | 10.9 (17.7) [0.5, 169.0] |
| N1 duration (min) | 46.6 (27.1) [7.0, 188.5] | 129.2 (58.4) [21.0, 317.0] |
| N2 duration (min) | 165.8 (50.4) [21.0, 289.0] | 123.4 (52.1) [0.0, 254.0] |
| N3 duration (min) | 83.3 (43.7) [1.5, 227.5] | 38.4 (30.9) [0.0, 139.0] |
| REM duration (min) | 78.2 (34.0) [0.0, 181.5] | 69.3 (29.4) [0.0, 170.5] |
| N1 percentage (%) | 13.4 (9.3) [2.8, 71.5] | 37.0 (18.0) [4.9, 100.0] |
| N2 percentage (%) | 44.2 (8.5) [20.5, 63.7] | 33.7 (12.2) [0.0, 58.0] |
| N3 percentage (%) | 22.4 (11.1) [0.9, 53.1] | 10.5 (8.2) [0.0, 36.1] |
| REM percentage (%) | 20.1 (6.6) [0.0, 38.5] | 18.8 (6.7) [0.0, 40.5] |
| ArI (events/h) | 16.2 (9.6) [3.2, 78.1] | 46.0 (18.8) [7.3, 116.6] |
| AHI (events/h) | N/A | 39.1 (19.6) [6.7, 103.0] |
| Sleep apnea severity |  |  |
| Mild (N, %) | N/A | 19 (8.1) |
| Moderate (N, %) | N/A | 69 (29.5) |
| Severe (N, %) | N/A | 146 (62.4) |
| Very severe (N, %) | N/A | 62 (26.5) |

Continuous variables are presented as mean (standard deviation) [min, max], and categorical variables are presented as count (percentage). TIB, time in bed; TST, total sleep time; SE, sleep efficiency; WASO, wake after sleep onset; SL, sleep latency; ArI, arousal index; AHI, apnea-hypopnea index. N1 duration is total time in N1 sleep (N2, N3, and REM durations defined similarly). N1 percentage is N1 duration as a proportion of TST (N2, N3, and REM percentages defined similarly).
Mild sleep apnea: $5 \leq \text{AHI} < 15$ ; moderate: $15 \leq \text{AHI} < 30$ ; severe: $\text{AHI} \geq 30$ ; very severe: $\text{AHI} \geq 50$ .

#### 2.2.1 Ethics Statement

This study was approved by the Ethics Committee of Hillside Clinic Jingumae (Approval Nos. SUG08869 and SUG08884). Written informed consent was obtained from all participants before data collection. All recordings were anonymized before analysis to prevent identification of individual participants.

#### 2.2.2 Sleep Lab Dataset

The Sleep Lab Dataset comprised 318 overnight recordings acquired at a sleep laboratory in Japan. PSG and wearable recordings were acquired simultaneously. Participants were paid volunteers recruited through general advertisement. Because recruitment was not for diagnostic purposes, this dataset should not be interpreted as a clinically confirmed healthy cohort. After data collection, sleep stages and arousal events were manually scored. Respiratory events were not scored in this dataset, and AHI was therefore unavailable. Eleven participants contributed multiple nights (2–3 nights), which were recorded on non-consecutive days. To prevent data from the same participant appearing in both training and evaluation sets, subject-level splitting was employed in the experimental design described below.

#### 2.2.3 Hospital Dataset

The Hospital Dataset comprised 234 overnight recordings acquired at Toranomon Hospital in Japan. PSG and wearable recordings were acquired simultaneously. Participants were individuals who underwent clinical assessment for sleep apnea. After data collection, sleep stages, arousal events, and respiratory events were manually scored. Sleep apnea severity, as measured by AHI, had a mean of 39.1 and a maximum of 103.0. For the present study, sleep apnea subtype was not classified diagnostically, and analyses were based on respiratory event scoring and AHI. All participants were unique individuals, with no repeated measurements. More than half of the Hospital Dataset (62.4%) had severe sleep apnea, defined as AHI ≥ 30. Approximately one-quarter (26.5%) met the criterion for very severe sleep apnea, defined as AHI ≥ 50, providing substantial representation of high-severity cases. In subsequent evaluations, when further subdividing high-severity cases by AHI, we used 50 events/h as the threshold. Although some prior studies define very severe sleep apnea using thresholds of 55 or 60 events/h [25, 26], there is currently no widely accepted threshold. Therefore, we selected the threshold to yield approximately comparable sample sizes across the subdivided groups.

#### 2.2.4 Comparison of the Two Datasets

The two datasets were used as comparison cohorts with different participant characteristics and PSG-derived sleep patterns, rather than as a severity-matched case-control contrast. Compared with the Sleep Lab Dataset, participants in the Hospital Dataset were older, more often male, and had higher body mass index (BMI). PSG-derived sleep parameters also differed, with the Hospital Dataset showing higher arousal index (ArI) and N1 duration and lower N3 duration and sleep efficiency (SE) than the Sleep Lab Dataset. Because respiratory events were not scored in the Sleep Lab Dataset, AHI was unavailable and sleep apnea severity could not be directly compared between the two datasets. Undiagnosed sleep-disordered breathing in the Sleep Lab Dataset could therefore not be excluded. Accordingly, the Sleep Lab Dataset was treated as a non-diagnostic, reference-like cohort with relatively less fragmented sleep on PSG-derived markers, rather than as a clinically confirmed healthy cohort.

### 2.3 Data Preprocessing and Feature Extraction

For sleep staging, we extracted input features from RRI and acceleration signals using the following preprocessing pipeline, with steps applied in the listed order.

1. **RRI outlier removal**: RRI values exceeding 3000 ms were excluded as physiologically implausible values, taking possible signal contamination or beat-detection errors into account. The corresponding positions were then left without an RRI value.
2. **Acceleration feature extraction**: For each of the three acceleration axes, the mean and standard deviation were computed from the raw acceleration waveform in consecutive non-overlapping 5-second windows, yielding six acceleration-derived features. Window start times were aligned to timestamps whose seconds were multiples of 5.
3. **Resampling**: The seven resulting features (RRI and six acceleration features) were resampled to 8 Hz. During resampling, values at 8-Hz target time points between neighboring observed samples were obtained by linear interpolation. For features observed over shorter data than the other features, the nearest observed value was carried forward/backward at the start and end so that all features shared aligned start and end times.
4. **Standardization**: The seven resampled features were linearly transformed to have zero mean and unit variance. Pre-specified feature-wise mean and standard deviation parameters, determined in advance from data independent of the present study datasets, were applied unchanged to both the Sleep Lab Dataset and the Hospital Dataset.

These acceleration features summarized the raw acceleration waveform in consecutive 5-second windows and simplified the input representation passed to the model.

### 2.4 Deep Learning Model

#### 2.4.1 Input Representation

Sleep staging was performed at a 30-second epoch resolution. For each epoch, we used a 32-second input window spanning the target 30-second epoch plus 1 second before and 1 second after. Thus, the input for each epoch had shape (7, 256), corresponding to 7 features sampled over 32 sec at 8 Hz.

#### 2.4.2 Network Architecture

Hybrid convolutional neural network (CNN) and recurrent neural network (RNN) architectures are widely used for physiological time-series classification and have been adopted in prior sleep staging studies [17, 27, 28]. We implemented such an architecture consisting of a 1-dimensional CNN encoder, an RNN module, and a classification head. This design was motivated by the dual temporal structure of sleep staging: (i) within-epoch patterns that reflect short-term autonomic and movement-related fluctuations, such as transient changes in RRI and acceleration around arousals; and (ii) across-epoch dynamics that reflect stage transitions and sleep architecture over minutes to tens of minutes. Accordingly, the CNN encoder was used to extract an epoch-level feature from the 32-second input window, while the RNN module was used to model temporal dependencies across consecutive epochs within a night.

An overview of the model architecture is provided in Table 2, and the detailed CNN architecture is shown in Table 3.

**Table 2:**
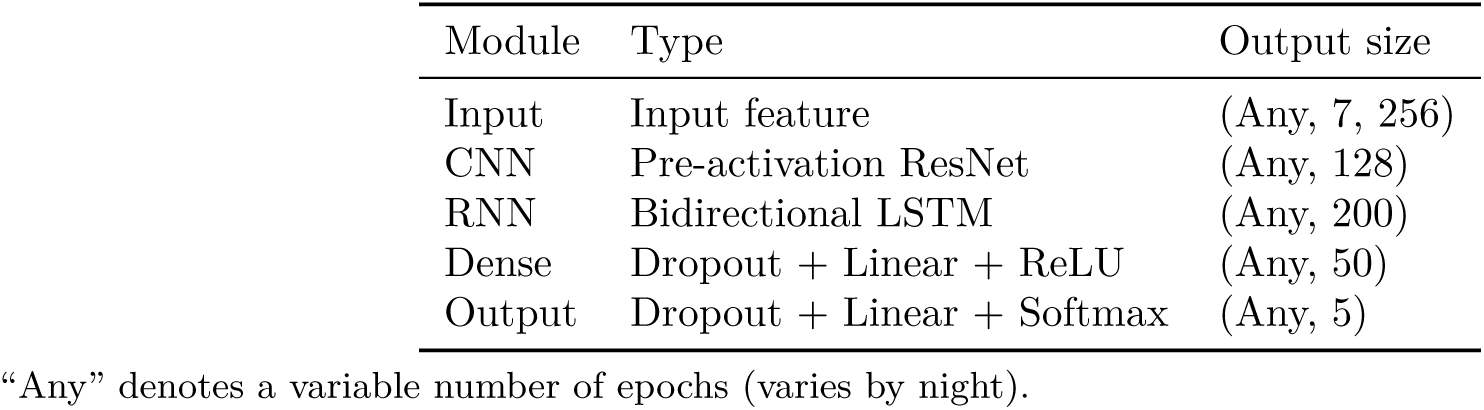
High-level architecture of the deep learning model.

**Table 3:**
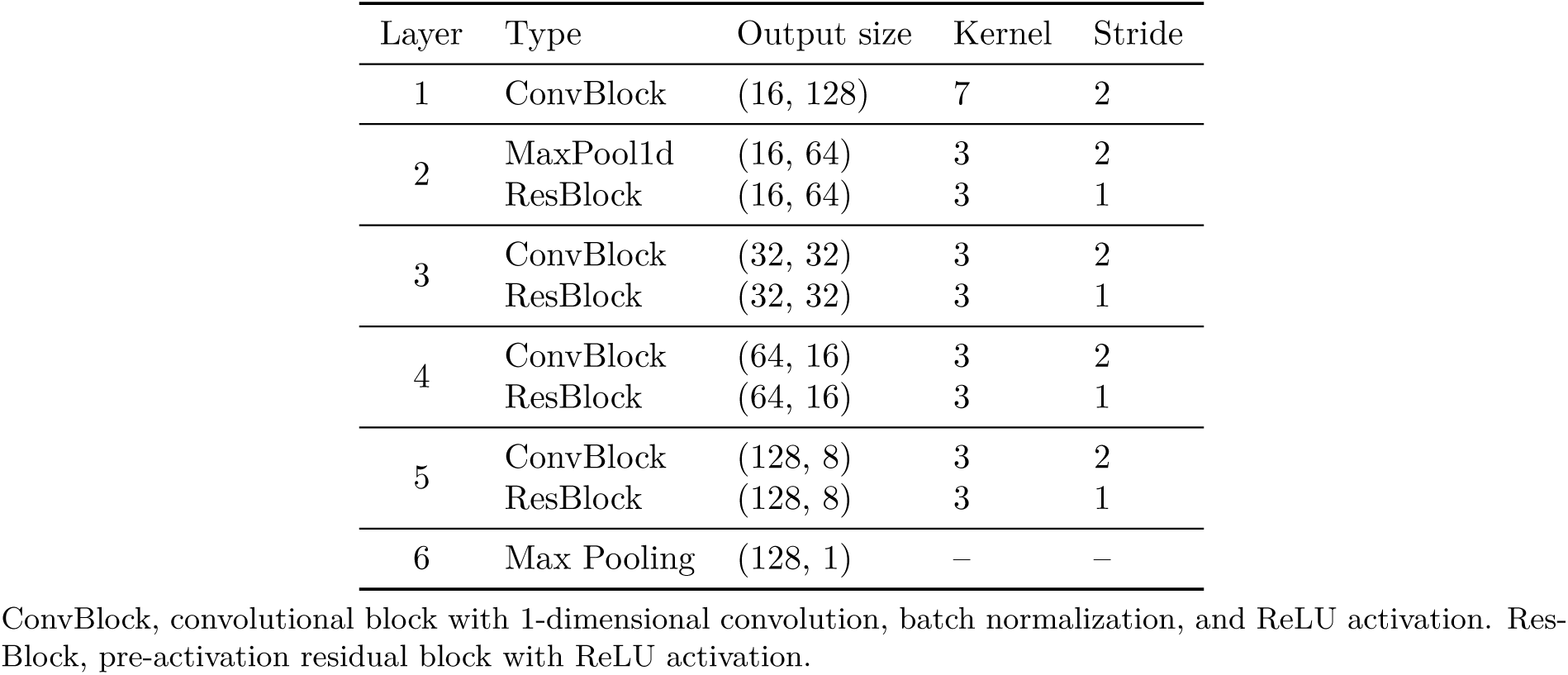
Detailed architecture of the CNN encoder (ResNet).

The CNN encoder was based on a ResNet-style architecture [29, 30] adapted to 1-dimensional features to extract an epoch-level feature. The RNN module was a 2-layer bidirectional long short-term memory (LSTM) network [31, 32] with 100 hidden units per direction, producing a 200-dimensional feature vector per epoch. The classification head comprised two fully connected dense layers that map a 200-dimensional input to 50 dimensions and then to 5 dimensions, with ReLU activations, followed by softmax to output predicted probabilities for the five sleep stages (Wake, N1, N2, N3, and REM). Dropout (rate 0.2) was applied within the RNN module and classification head.

#### 2.4.3 Training

The model was trained for 5-stage classification. During training, 10% of the training set was used as a validation set for hyperparameter tuning and early stopping. Training was performed for up to 300 training epochs with early stopping. Because recording duration varied across recordings, sequences within each mini-batch were zero-padded at the end using post-padding to match the maximum number of epochs in the mini-batch. Selected hyperparameters are summarized in Table 4.

**Table 4:** Training hyperparameters.

| Parameter | Value |
| --- | --- |
| Optimizer | AdamW |
| Learning rate | 0.001 |
| Loss function | Focal loss ($\gamma = 2.0$) |
| Batch size | 32 |
| Max training epochs | 300 |

Training required approximately 1 minute per training epoch on an AWS EC2 g4dn instance with an NVIDIA T4 GPU.

#### 2.4.4 Output and Stage Mapping

For each epoch, the predicted sleep stage was defined as the class with the highest predicted probability. Because 4-stage classification (Wake, Light [N1+N2], Deep [N3], REM) is commonly reported in wearable sleep staging studies, we report evaluation results under both 5-stage and 4-stage staging schemes. For 4-stage classification, we first defined the 5-stage predicted stage as the stage with the highest predicted probability, and then mapped that predicted stage by collapsing N1 and N2 into the Light class.

### 2.5 Experimental Design

In all evaluations, PSG-based manual scoring by certified technicians served as the reference standard. We conducted three experiments:

#### 2.5.1 Experiment 1: Baseline Performance

To assess baseline performance across the full dataset, we performed 4-fold cross-validation on the combined dataset of 552 recordings. The Sleep Lab Dataset and Hospital Dataset were each randomly divided into four folds, and the corresponding folds from the two datasets were then combined to form four combined folds of 138 recordings each. Subject-level grouping was used in the Sleep Lab Dataset to ensure that multiple nights from the same participant were assigned to the same fold. For each fold, the model was trained on the remaining three folds and evaluated on the hold-out fold. We evaluated epoch-wise sleep staging under both 5-stage and 4-stage schemes and summarized epoch-wise performance metrics as the mean and standard deviation across folds. We also aggregated the cross-validation predictions across folds to compute confusion matrices and derived sleep parameters from the predicted stages for night-level agreement analyses.

#### 2.5.2 Experiment 2: Night-level Performance by AHI Group

Using the predictions obtained in Sec. 2.5.1, we examined whether night-level performance varied across AHI groups in the Hospital Dataset. For analyses by AHI group, we categorized AHI as mild (5 ≤ AHI < 15), moderate (15 ≤ AHI < 30), severe (30 ≤ AHI < 50), and very severe (AHI ≥ 50). Performance for 5-stage sleep staging was assessed using Cohen’s kappa. To provide supplementary exploratory context, we also summarized associations of performance with demographic variables and sleep parameters.

#### 2.5.3 Experiment 3: Training-Set Composition and Evaluation in Very Severe Sleep Apnea

To evaluate how training-set composition affected performance in very severe sleep apnea, we conducted an ablation experiment while fixing the training sample size and evaluation set. The evaluation set comprised all Hospital Dataset recordings with AHI ≥ 50 (*N* = 62). We compared the baseline model from Sec. 2.5.1 with an ablation model trained on an alternative training set. For ablation training, we fixed the sample size to *N* = 414 recordings, matching one training split in the 4-fold baseline setting, and constructed the training set using all Sleep Lab Dataset recordings (*N* = 318) plus the lowest-AHI Hospital Dataset recordings until the target size was reached (*N* = 96), which corresponded to AHI < 32. This ablation training set included substantially fewer high-AHI cases while keeping the total number of training cases unchanged. Performance metrics from the baseline and ablation models were compared as paired observations for each evaluation recording.

## 3 Results

### 3.1 Baseline Model Performance

#### 3.1.1 Epoch-wise Sleep Staging Performance

We evaluated epoch-wise sleep staging performance using accuracy, Cohen’s kappa, and F1-score. Results are presented as mean and standard deviation across 4-fold cross-validation. Overall performance is summarized in Table 5 for 5-stage and Table 6 for 4-stage. For 5-stage classification, the Hospital Dataset showed lower kappa than the Sleep Lab Dataset, with a between-dataset gap of 0.140. Stage-specific performance showed a consistent pattern across datasets: N1 remained the most difficult stage to classify, whereas Wake and REM were classified more accurately. For 4-stage classification, the between-dataset gap narrowed to 0.107.

**Table 5:** Overall performance for 5-stage sleep staging.

| Dataset | Accuracy | Kappa | F1-score |  |  |  |  |  |
| --- | --- | --- | --- | --- | --- | --- | --- | --- |
|  |  |  | Macro | Wake | N1 | N2 | N3 | REM |
| Sleep Lab | 0.694 | 0.586 | 0.667 | 0.737 | 0.402 | 0.714 | 0.691 | 0.788 |
|  | (0.017) | (0.023) | (0.019) | (0.040) | (0.021) | (0.013) | (0.023) | (0.021) |
| Hospital | 0.574 | 0.446 | 0.580 | 0.666 | 0.500 | 0.557 | 0.525 | 0.651 |
|  | (0.005) | (0.006) | (0.009) | (0.016) | (0.020) | (0.012) | (0.041) | (0.023) |

Performance metrics are presented as mean (standard deviation) across 4-fold cross-validation.

**Table 6:** Overall performance for 4-stage sleep staging.

| Dataset | Accuracy | Kappa | F1-score |  |  |  |  |
| --- | --- | --- | --- | --- | --- | --- | --- |
|  |  |  | Macro | Wake | Light | Deep | REM |
| Sleep Lab | 0.760<br>(0.013) | 0.632<br>(0.022) | 0.749<br>(0.019) | 0.737<br>(0.040) | 0.778<br>(0.012) | 0.691<br>(0.023) | 0.788<br>(0.021) |
| Hospital | 0.713<br>(0.009) | 0.525<br>(0.010) | 0.653<br>(0.014) | 0.666<br>(0.016) | 0.771<br>(0.009) | 0.525<br>(0.041) | 0.651<br>(0.023) |

Performance metrics are presented as mean (standard deviation) across 4-fold cross-validation. Light combines N1 and N2; Deep corresponds to N3.

#### 3.1.2 Sleep Staging Error Structure (Confusion Matrices)

We aggregated epoch-wise predictions across the cross-validation folds to compute confusion matrices for 5-stage and 4-stage sleep staging. The results are shown in Fig. 2 and Fig. 3. In 5-stage classification, the most frequent errors in both datasets involved confusions around N2, in particular misclassifying N1 and N3 as N2. N1 was more often misclassified as N2 than as Wake in both datasets, and N3 was primarily misclassified as N2. Beyond these N2-centered errors, misclassification was largely confined to adjacent stages, whereas non-adjacent confusions were relatively uncommon. Compared with Sleep Lab Dataset, Hospital Dataset showed overall increased misclassification and more pronounced confusions between adjacent stages, including more N2 epochs classified as N1 and more Wake epochs classified as N1.

**Figure 2:**
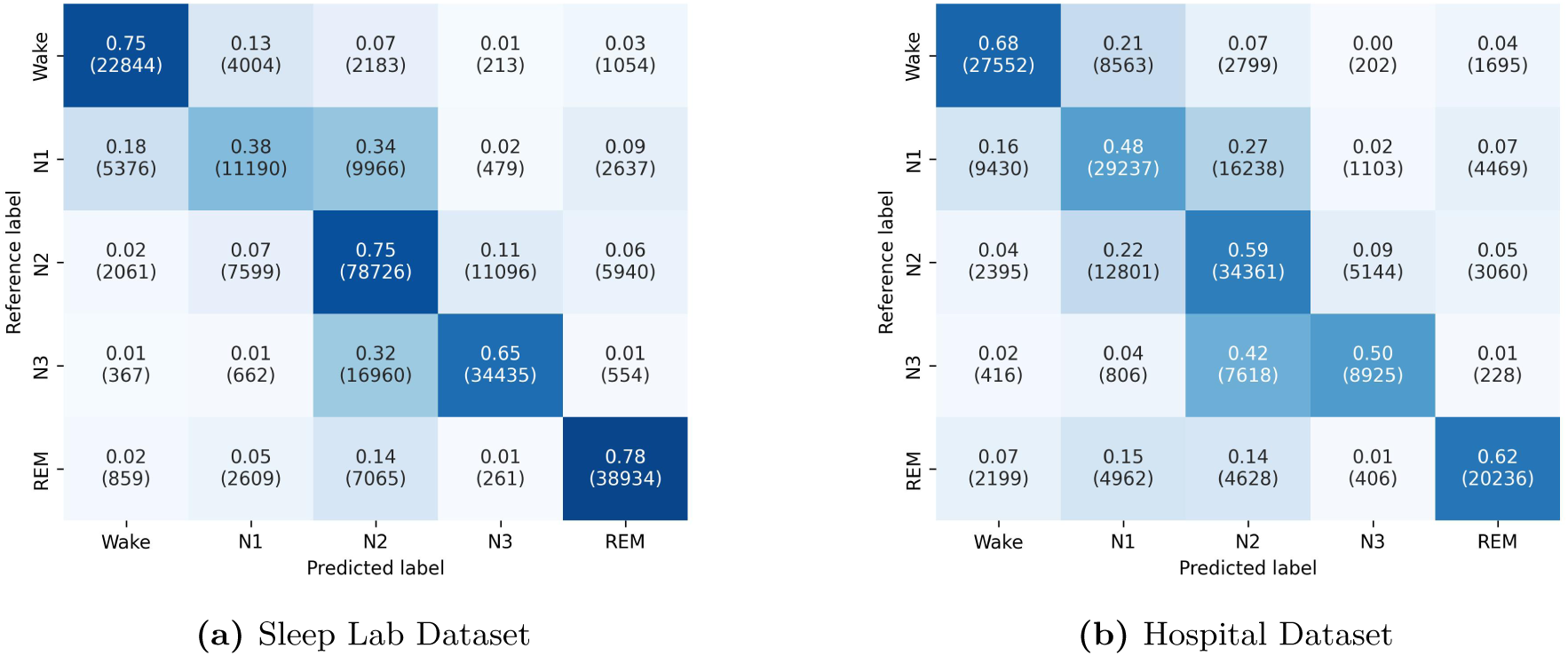
Confusion matrices for 5-stage sleep staging. Results are shown for (a) Sleep Lab Dataset and (b) Hospital Dataset. Rows indicate the reference (PSG) label and columns indicate the predicted label. Each cell reports the row-normalized proportion of epochs with the epoch count in parentheses. Color intensity reflects the proportion.

**Figure 3:**
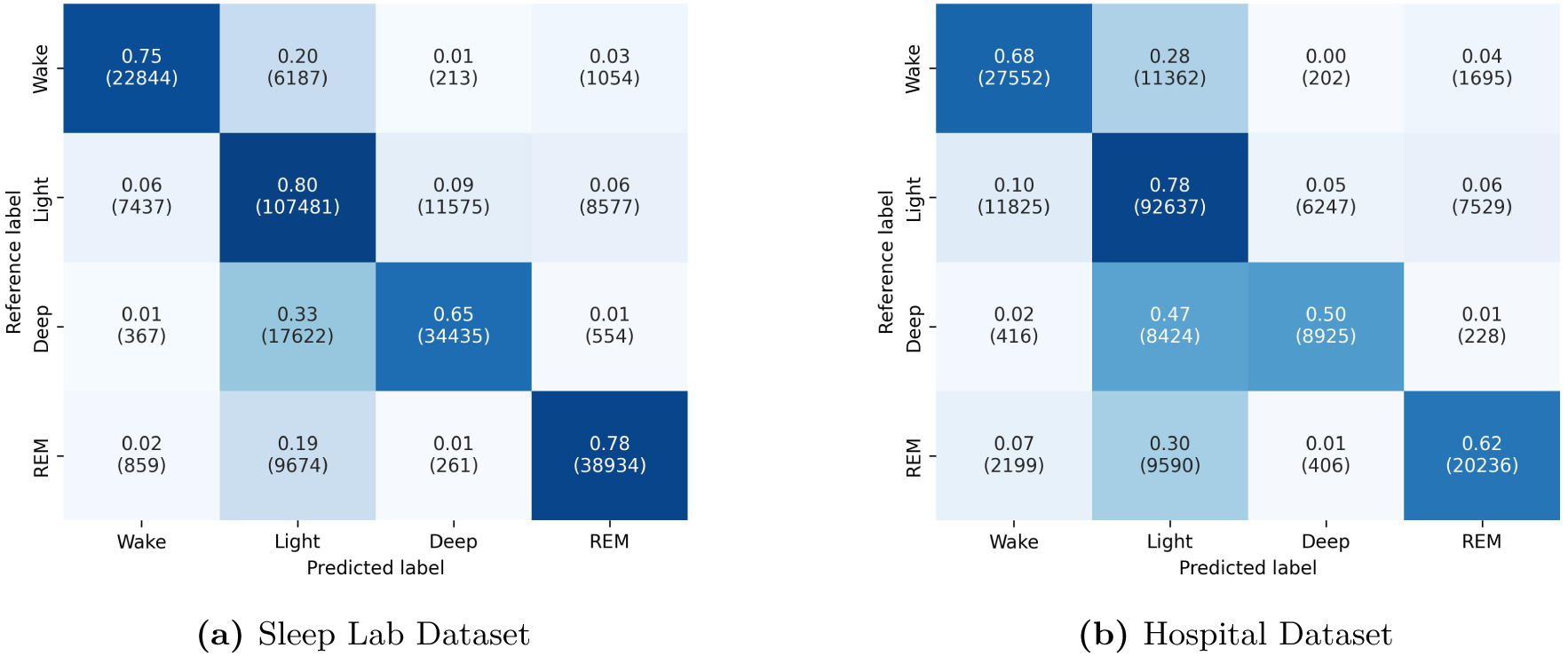
Confusion matrices for 4-stage sleep staging. Results are shown for (a) Sleep Lab Dataset and (b) Hospital Dataset. Rows indicate the reference (PSG) label and columns indicate the predicted label. Each cell reports the row-normalized proportion of epochs with the epoch count in parentheses. Color intensity reflects the proportion. Light combines N1 and N2, and Deep corresponds to N3.

In 4-stage classification, N1 and N2 are combined into Light, so the N1–N2 confusions observed in 5-stage classification are subsumed within the Light class. Accordingly, the remaining errors were dominated by confusions involving Light versus other stages, particularly Light–Deep confusions. The Hospital Dataset also showed more Deep epochs misclassified as Light. Overall, non-adjacent confusions such as Wake versus Deep or REM were uncommon, typically below 7%.

#### 3.1.3 Night-level Sleep Parameter Agreement

We computed sleep parameters from the predicted stages and compared them with the PSG reference. Target parameters were total sleep time (TST), sleep efficiency (SE), wake after sleep onset (WASO), sleep latency (SL), and the durations of N1, N2, N3, and REM. For each parameter, we report the mean and standard deviation of the PSG reference and device-predicted values. We also report bias, defined as device minus reference, together with limits of agreement (LoA) and mean absolute error (MAE).

Overall, MAE tended to be smaller in the Sleep Lab Dataset than in the Hospital Dataset, and LoA were generally wider in the Hospital Dataset. Table 7 summarizes these comparisons. Among the stage-duration parameters in the Hospital Dataset, N1 duration had the widest LoA, and N2 duration had the largest MAE. Systematic bias patterns were observed: N2 duration and SL showed positive bias in both datasets, whereas N1 and N3 duration showed negative bias. For TST and SE, the absolute bias was comparatively small, less than 1% of the reference mean in both datasets.

**Table 7:** Sleep parameter estimation performance.

| Parameter | Dataset | Mean (SD) |  | Bias | LoA | MAE |
| --- | --- | --- | --- | --- | --- | --- |
|  |  | Reference | Device |  |  |  |
| TST | Sleep Lab | 373.9 (87.8) | 372.0 (86.0) | -1.9 | [-60.1, 56.3] | 16.2 |
|  | Hospital | 360.4 (56.5) | 357.9 (70.0) | -2.5 | [-119.5, 114.4] | 35.6 |
| SE | Sleep Lab | 88.7 (12.5) | 88.4 (12.5) | -0.3 | [-14.3, 13.7] | 3.9 |
|  | Hospital | 80.7 (12.5) | 80.1 (15.4) | -0.6 | [-26.7, 25.5] | 7.9 |
| WASO | Sleep Lab | 39.5 (50.1) | 34.7 (45.1) | -4.8 | [-66.8, 57.3] | 16.2 |
|  | Hospital | 71.5 (53.5) | 71.6 (59.4) | 0.1 | [-94.5, 94.7] | 29.4 |
| SL | Sleep Lab | 5.7 (10.7) | 11.7 (25.6) | 5.9 | [-38.2, 50.1] | 8.3 |
|  | Hospital | 10.9 (17.7) | 13.4 (20.5) | 2.3 | [-34.6, 39.6] | 9.4 |
| N1 duration | Sleep Lab | 46.6 (27.1) | 41.0 (31.7) | -5.6 | [-55.0, 43.8] | 18.1 |
|  | Hospital | 129.2 (58.4) | 120.4 (64.7) | -8.8 | [-129.7, 112.2] | 46.2 |
| N2 duration | Sleep Lab | 165.8 (50.4) | 180.7 (54.8) | 14.9 | [-68.6, 98.4] | 34.8 |
|  | Hospital | 123.4 (52.1) | 140.3 (60.0) | 16.8 | [-101.5, 135.1] | 48.2 |
| N3 duration | Sleep Lab | 83.3 (43.7) | 73.1 (32.0) | -10.2 | [-89.4, 69.0] | 31.6 |
|  | Hospital | 38.4 (30.9) | 33.7 (29.3) | -4.7 | [-71.1, 61.7] | 24.6 |
| REM duration | Sleep Lab | 78.2 (34.0) | 77.2 (32.3) | -1.0 | [-39.3, 37.4] | 14.5 |
|  | Hospital | 69.3 (29.4) | 63.4 (31.1) | -5.8 | [-64.6, 52.8] | 22.5 |

Reference values are from PSG; device values are from stage predictions derived from wearable recordings. Reference and device values are presented as mean (standard deviation) across recordings. Bias is the mean difference (device minus reference). LoA denotes the limits of agreement and is calculated as bias ± 1.96 times the standard deviation. Durations are in minutes and SE is in percentage points.
TST, total sleep time; SE, sleep efficiency; WASO, wake after sleep onset; SL, sleep latency; MAE, mean absolute error.

### 3.2 Night-level Performance by AHI Group

Fig. 4 shows the overall Sleep Lab Dataset distribution for context and the grouped comparison across AHI groups within the Hospital Dataset. In the Hospital Dataset, night-level Cohen’s kappa for 5-stage sleep staging shifted downward from lower to higher AHI groups. The median kappa difference between the mild and very severe groups was close to 0.2. Supplementary analyses likewise showed a negative association between AHI and sleep staging performance (Pearson *r* = −0.360). Detailed results are provided in Supplementary Table S1 and Supplementary Fig. S3. Additional scatter plots for demographic variables and other sleep parameters are provided in Supplementary Sec. A.1.

**Figure 4:**
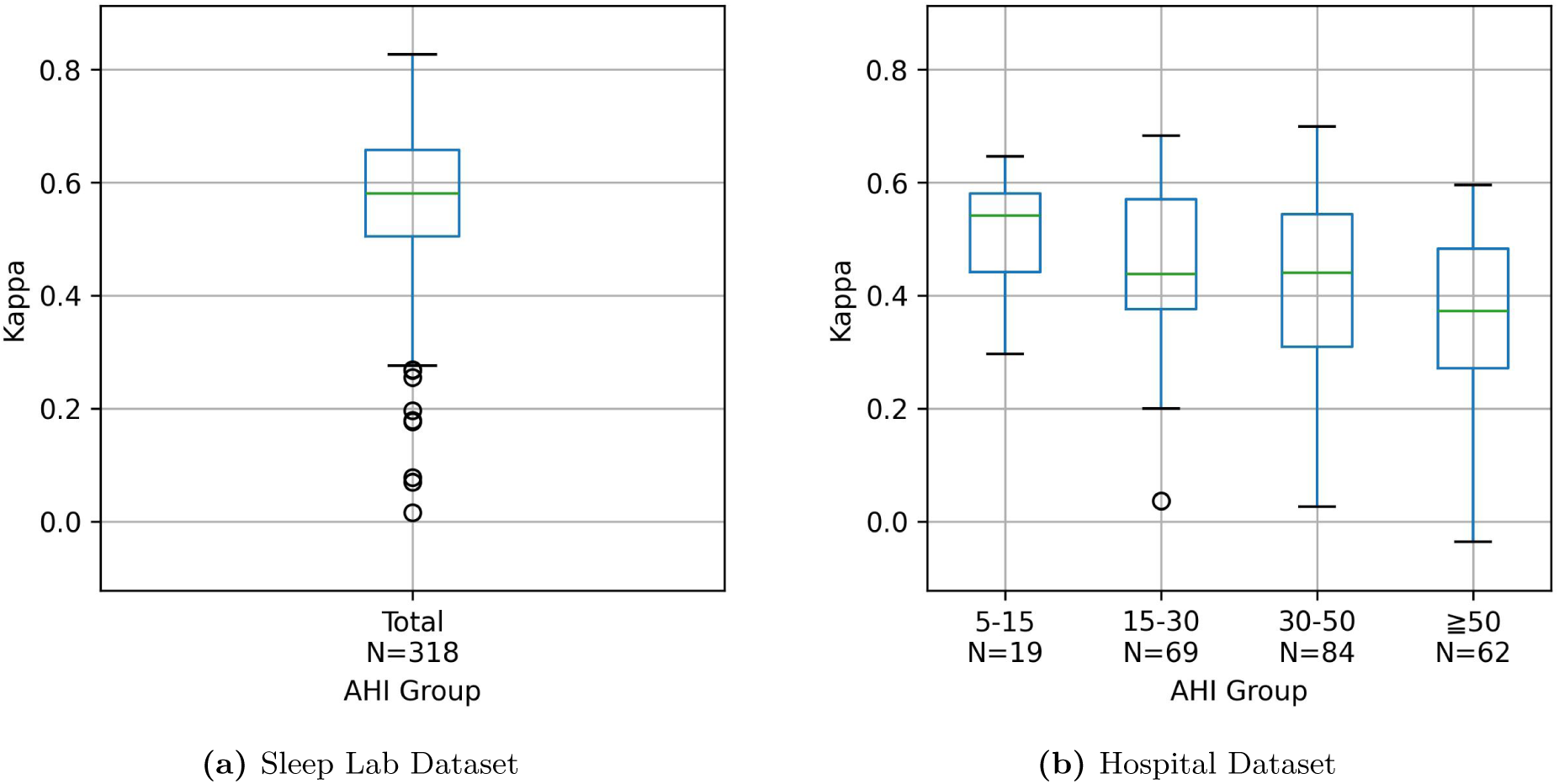
Sleep staging performance by dataset and AHI group. The y-axis shows night-level Cohen’s kappa for 5-stage sleep staging. Panel (a) shows the overall Sleep Lab Dataset distribution, and panel (b) shows Hospital Dataset results by AHI group. A label *x*–*y* denotes the range *x ≤* AHI < *y* events/h (e.g., 15–30 covers 15 ≤ AHI < 30). Boxes indicate the interquartile range with the median shown as a horizontal line, whiskers extend to 1.5 times the interquartile range, and points indicate outliers.

### 3.3 Effect of Training-set Composition in Very Severe Sleep Apnea

A paired scatter plot of night-level kappa values is shown in Fig. 5. Summaries for kappa and macro F1 are reported in Table 8. Compared with baseline training, the ablation setting showed a clear downward shift in night-level kappa values, with both kappa and macro F1 lower by about 5–6 points.

**Figure 5:**
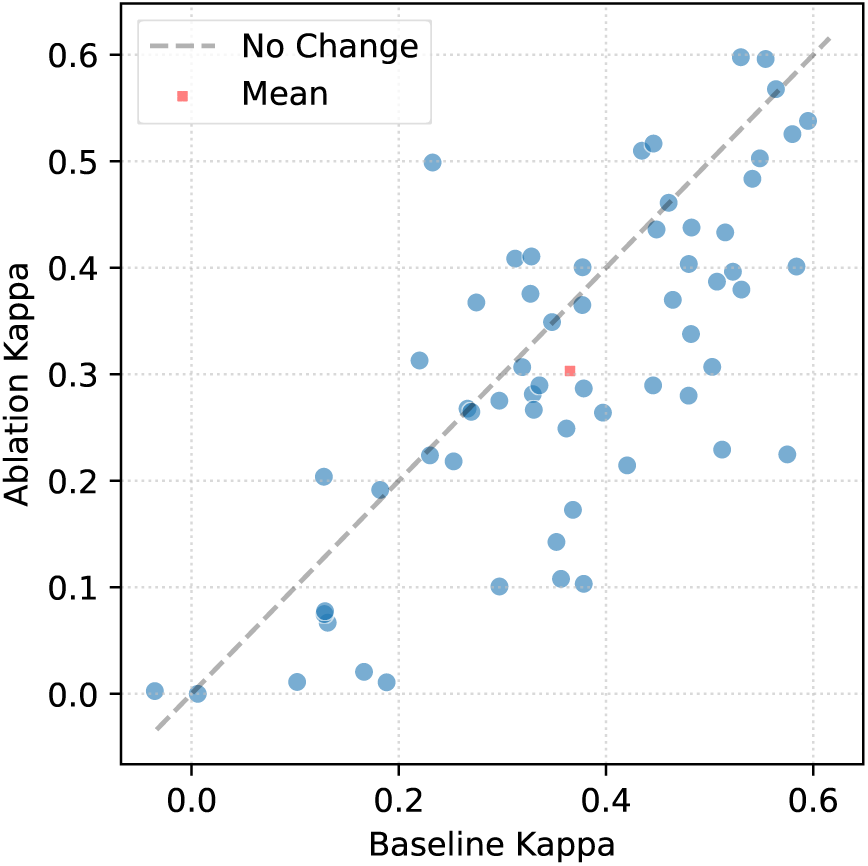
Paired scatter plot of night-level kappa values for 5-stage sleep staging in Hospital Dataset recordings with AHI ≥ 50 (*N* = 62). The x-axis shows the baseline kappa and the y-axis shows the ablation kappa. Baseline denotes the model trained on the full combined Sleep Lab Dataset and Hospital Dataset recordings. Ablation denotes the model trained on the same number of recordings, using all Sleep Lab Dataset recordings and Hospital Dataset recordings with AHI < 32. Each point represents one recording. The gray dashed diagonal indicates no change between models, and the red marker indicates the mean pair of kappa values.

**Table 8:** Impact of training data composition on performance in very severe sleep apnea.

| Metric | Mean |  | Paired difference |  |
| --- | --- | --- | --- | --- |
|  | Baseline | Ablation | Estimate | 95% CI |
| Kappa | 0.365 | 0.303 | −0.062 | [−0.090, −0.035] |
| Macro F1 | 0.458 | 0.408 | −0.049 | [−0.072, −0.029] |

All values are based on 5-stage sleep staging evaluated in Hospital Dataset recordings with $\text{AHI} \geq 50$ ( $N = 62$ ). Values in the Baseline and Ablation columns are means across evaluation recordings. Baseline denotes the cross-validation model trained on the full combined Sleep Lab Dataset and Hospital Dataset recordings. Ablation denotes the model trained on the same number of recordings as the baseline training split, using all Sleep Lab Dataset recordings and Hospital Dataset recordings with $\text{AHI} < 32$ . Paired differences were calculated for each recording as $\text{Ablation} - \text{Baseline}$ . The estimate is the mean of the night-level paired differences, with 95% confidence intervals (CI) estimated by the bias-corrected and accelerated (BCa) bootstrap method with 10,000 resamples.

## 4 Discussion

### 4.1 Summary of Main Findings

This study evaluated wrist-worn wearable sleep staging in 552 overnight recordings from two complementary cohorts, a non-diagnostic sleep-laboratory dataset and a clinical sleep apnea dataset that included a sizable very severe subgroup (*N* = 62). For 5-stage classification, Cohen’s kappa was 0.586 in the Sleep Lab Dataset and 0.446 in the Hospital Dataset. Under 4-stage staging, the between-dataset gap narrowed, with Cohen’s kappa of 0.632 in the Sleep Lab Dataset and 0.525 in the Hospital Dataset, after collapsing N1 and N2 into the Light class. Sleep parameter estimation in the Hospital Dataset showed wider LoAs and larger MAEs than in the Sleep Lab Dataset. Within the Hospital Dataset, night-level Cohen’s kappa declined as AHI severity increased, with the median kappa differing by about 0.2 between the mild and very severe groups. We also compared the baseline model with an ablation model trained on the same number of recordings but with proportionally fewer high-AHI cases. In the very severe subgroup, the ablation model produced lower kappa than the baseline.

### 4.2 Comparison with Prior Studies

Many wearable sleep staging studies have been validated primarily in healthy or general populations, although some prior studies have included participants with sleep apnea [22, 23]. However, evidence remains limited for studies that specifically evaluated performance in a sizable subgroup with very severe sleep apnea, and prior sleep apnea studies often did not report results separately for severe or very severe subgroups. Against this background, our wrist-worn wearable study adds large-scale validation in a hospital dataset that included participants with very severe sleep apnea. For quantitative comparison, Table 9 prioritizes representative recent studies with staging granularity and sensor configurations closer to ours. We did not identify recent watch-based studies reporting 5-stage sleep staging with PPG and accelerometer. Therefore, for 5-stage reference, it also includes representative studies using PPG derived from PSG recordings. For 4-stage classification, performance in the Sleep Lab Dataset was comparable to prior watch studies, whereas performance in the Hospital Dataset was slightly lower. For 5-stage classification, a similar pattern was observed. In this case, performance in the Sleep Lab Dataset fell within the range reported in the literature, whereas performance in the Hospital Dataset was below that range. The lower performance of the Hospital Dataset relative to both the prior watch-based studies and the PSG-PPG comparison studies is consistent with the difference in AHI distribution. Specifically, the Hospital Dataset was shifted toward higher AHI values than the cohorts reported in the comparison studies, and in our data Cohen’s kappa decreased with increasing AHI. A similar trend has been reported in a recent watch-based study [20], in which AHI was negatively correlated with sleep-staging performance measured by balanced accuracy (Pearson *r* = −0.31, *p <* 0.001).

**Table 9:** Comparison of sleep staging performance with prior studies.

| Study | Year | Device | Sensor | N | AHI | Stages | Kappa |
| --- | --- | --- | --- | --- | --- | --- | --- |
| Fonseca et al. [17] | 2023 | Watch | PPG+ACC | 394 | N/A | 4 | 0.64 |
| Liu et al. [19] | 2023 | Watch | PPG+ACC | 75 | 2.3(3.1) | 4 | 0.55 |
| Silva et al. [20] | 2024 | Watch | PPG+ACC | 586 | 23.5(25.6) | 4 | 0.56 |
| Constantin et al. [21] | 2025 | Watch | PPG | 68 | 4.9[2.6, 14.0] | 4 | 0.66 |
| Korkalainen et al. [27] | 2020 | PSG | PPG | 89 | 16.8*[6.5, 33.2] | 5/4 | 0.51/0.54 |
| Huttunen et al. [28] | 2021 | PSG | PPG | 175 | 19.9*[8.2, 41.2] | 5/4 | 0.60/0.64 |
| **Ours (Sleep Lab)** | 2026 | Watch | PPG+ACC | 318 | N/A | 5/4 | 0.59/0.63 |
| **Ours (Hospital)** | 2026 | Watch | PPG+ACC | 234 | 39.1(19.6)[24.2, 51.5] | 5/4 | 0.45/0.53 |

Device: recording platform (wrist watch vs. laboratory PSG); Sensor: signal modalities used; $N$ : dataset size; AHI: apnea-hypopnea index; Stages: sleep staging scheme (4 or 5 classes); Kappa: Cohen’s kappa for overall agreement; PPG: photoplethysmography; ACC: accelerometry.
AHI is reported as mean (standard deviation) [Q1, Q3], where [Q1, Q3] denotes the interquartile range. If a study did not report the standard deviation or the interquartile range, those values are omitted. If no AHI summary was reported, the cell reads N/A. Entries such as 5/4 in the Stages column mean both five- and four-stage evaluations were reported.
\* The study reported the median AHI rather than the mean.

### 4.3 Sources of Error and Performance Degradation

Manual PSG scoring has non-negligible inter-scorer variability, which reduces the attainable performance ceiling and adds noise to both training and evaluation [33–35]. This variability is particularly pronounced for N1: prior work has reported overall inter-scorer agreement typically around kappa 0.7–0.8 and macro F1 score 0.7–0.8, whereas agreement for N1 is substantially lower, with kappa around 0.2–0.5 and F1 score around 0.4–0.5 [33–35]. These findings suggest that the N1–N2 boundary is intrinsically uncertain even in the reference labels, constraining the attainable performance ceiling. This interpretation is consistent with the frequent N1/N2 confusion in our results and with the improved performance observed when N1 and N2 were collapsed into the Light class in 4-stage staging. In sleep apnea cohorts, where scorer agreement may further decrease [35] and frequent arousals repeatedly drive transitions into lighter sleep, this source of difficulty may be further amplified. Furthermore, systematic confusions between adjacent stages can propagate to sleep parameter estimation, contributing to stage-duration biases such as overestimation of N2 duration and underestimation of N3 duration. These patterns are summarized in Table 7.

In these validation results, night-level staging difficulty in this clinical cohort primarily tracked AHI severity. Additional analyses of non-AHI variables were included as descriptive exploratory context rather than as tests of independent effects. These supplementary results included correlation summaries and omnibus grouped comparisons for demographic variables and sleep parameters other than AHI. Details are provided in Supplementary Table S1, Supplementary Table S2, and Supplementary Figs. S13–S24. In the supplementary analyses, ArI, N1 duration, and N3 duration tended to show the more consistent association patterns across both datasets, whereas TST, SE, WASO, and SL showed weaker or less consistent patterns.

Reflective wrist PPG is sensitive to motion artifacts, peripheral perfusion, and individual characteristics such as age, and degraded signal quality may lead to night-level failures [36]. In our data, a small number of nights exhibited extreme errors, including kappa < 0.1. These cases may reflect signal-related failures rather than typical model behavior. Examples of such nights can also be seen in Supplementary Sec. A.1.

### 4.4 Limitations

The two datasets were intentionally complementary rather than severity-matched. The Sleep Lab Dataset served as a non-diagnostic, reference-like cohort, whereas the Hospital Dataset comprised individuals undergoing clinical assessment for sleep apnea. Because respiratory events were not scored in the Sleep Lab Dataset, AHI was unavailable for that cohort, precluding direct comparison of severity distributions between datasets. In addition, undiagnosed sleep-disordered breathing could not be ruled out in the Sleep Lab Dataset. Therefore, the observed performance gap between the Sleep Lab Dataset and Hospital Dataset could not be used to determine how strongly sleep apnea severity contributed to the between-dataset difference. The gap may also reflect other cohort differences, including differences in input and label distributions.

As discussed in Sec. 4.3, the PSG reference labels are not error-free. Such label noise can both obscure true model improvements and complicate stage-specific interpretation when model predictions disagree with uncertain reference labels.

This study included only Japanese participants and focused on sleep apnea. Generalization to other populations, ethnicities, and other sleep disorders remains unverified [37, 38]. Differences in physiology, comorbidity profiles, and sleep apnea phenotypes across populations may affect both wearable signal characteristics and model performance. Therefore, the generalizability of these findings to heterogeneous clinical settings remains uncertain.

This study used a single device under simultaneous PSG recording conditions. Transferability to other wearable devices and validity under real-world home monitoring, including multi-night use without first-night effects, remain unverified. Real-world home use may involve different wearing behaviors, ambient conditions, and missing-data patterns that could degrade performance relative to controlled recordings.

### 4.5 Clinical and Practical Implications

In clinical sleep apnea cohorts, reporting 4-stage staging alongside 5-stage staging may be practically useful because the N1/N2 boundary is challenging and a coarser staging scheme can provide more stable summaries of sleep architecture. For applications requiring finer staging, 5-stage staging remains appropriate and informative, provided that uncertainty around the N1/N2 boundary is taken into account. For example, presenting 5-stage results together with confidence measures or stage-specific reliability indicators can help users avoid over-interpreting N1-related estimates.

Sleep parameter estimation in the Hospital Dataset generally showed larger MAEs and wider LoAs than in the Sleep Lab Dataset, suggesting that single-night estimates should be interpreted cautiously in this more challenging clinical cohort. Repeated measurements across multiple nights may yield more reliable assessments.

When training used the same number of recordings but included all Sleep Lab Dataset recordings and proportionally more lower-AHI Hospital Dataset recordings, performance decreased in recordings with very severe sleep apnea. This suggests that, for model development targeting severe sleep apnea populations, performance may benefit from development datasets that include sufficient representation of participants with severe or very severe disease rather than relying mainly on sleep-laboratory recordings or lower-AHI cases.

### 4.6 Future Directions

To establish external validity in real-world settings, multi-center validation in home environments over multiple nights is needed to assess generalizability and to mitigate potential first-night effects. Such studies should include diverse participant characteristics and usage conditions, and report not only average accuracy but also failure modes and between-night variability.

A clearer evaluation of the effect of AHI on night-level performance will require accounting for other sleep parameters and demographic variables. This would be facilitated by datasets covering a broader spectrum of sleep apnea severity, particularly through the inclusion of individuals without sleep apnea.

For home-monitoring applications, incorporating signal quality indices and prediction uncertainty estimates could enable the flagging of low-confidence nights and support more robust interpretation. In practice, reliability cues could be used to trigger re-measurement, down-weight unreliable nights in longitudinal summaries, and communicate uncertainty to clinicians and end users. It will also be important to investigate whether training or inference strategies tailored to clinically severe and unstable sleep patterns can reduce performance degradation in challenging nights.

To leverage public datasets, pretraining or transfer learning using large-scale resources such as SHHS [39] and MESA [40] could improve robustness and generalization across diverse populations. Further work could evaluate how much additional training on data from clinical cohorts such as severe sleep apnea is required, and whether this approach reduces performance degradation when models are applied to clinically severe populations.

## 5 Conclusion

We evaluated wrist-worn wearable sleep staging across a broad sleep apnea severity spectrum, including a sizable very severe subgroup. This study indicates that sleep apnea severity is an important consideration both when evaluating wearable sleep staging performance and when developing models for clinical sleep apnea populations. Staging granularity may also be selected according to the intended use, balancing broad nightly summaries with fine-grained stage-level interpretation. Future validation in multi-night home settings and broader, more diverse cohorts will be important for practical deployment.

## Author Contributions

**Sho Ogaki:** conceptualization, formal analysis, investigation, methodology, project administration, software, validation, visualization, writing – original draft, writing – review and editing. **Michiru Kaneda:** conceptualization, investigation, methodology, writing – review and editing. **Tomoyuki Nohara:** conceptualization, investigation, methodology, writing – review and editing. **Syuhei Fujita:** conceptualization, data curation, investigation, resources, writing – review and editing. **Naoshi Osako:** conceptualization, data curation, funding acquisition, investigation, project administration, resources, writing – review and editing. **Tomoko Yagi:** conceptualization, data curation, investigation, resources, writing – review and editing. **Yasuhiro Tomita:** data curation, resources, writing – review and editing. **Takanori Ogata:** conceptualization, funding acquisition, investigation, methodology, project administration, writing – review and editing.

## Data Availability

The data underlying this study are not publicly available due to ethical and privacy considerations related to human participant data.

Access to the data may be considered upon reasonable request to the corresponding author, subject to appropriate approvals.

## Acknowledgments

The authors thank Dr. Koji Ode, Dr. Akifumi Kishi, and Dr. Hiroki Ueda of the University of Tokyo for their constructive discussions and valuable insights throughout this study. Natural language processing tools were used to assist with English-language editing.

## Disclosure Statements

Financial Disclosure: This study was funded by ACCELStars, Inc., the developer of the wrist-worn wearable device evaluated in this study. SO, MK, TN, SF, NO, and TO are affiliated with the funder, and all other authors have financial relationships with the funder. Support took the form of salaries and fees for authors. Authors affiliated with the funder were involved in study design, data collection, analysis, and manuscript preparation, as described in the Author Contributions section.

## Non-financial Disclosure

None.

Preprint repositories: This manuscript is available as a preprint on medRxiv (https://doi.org/10.64898/2026.04.09.26350266).

## Notes

### Competing Interest Statement

SO, MK, TN, SF, NO, and TO are affiliated with ACCELStars, Inc.
All remaining authors have financial relationships with ACCELStars, Inc.

### Author Declarations

Ethics Committee of Hillside Clinic Jingumae gave ethical approval for this work (Approval Nos. SUG08869 and SUG08884).

### Summary of Updates

Title revised. Abstract changed from structured to unstructured format; keywords updated. Statement of Significance removed; Author Contributions section added. Disclosure statement revised to describe the funder's role. Text revised throughout for clarity. Supplemental material moved to a separate file. No changes to data, analyses, or results.

## References

1. Walker MP. The role of sleep in cognition and emotion. Ann. N. Y. Acad. Sci. 2009;1156(1):168–197. doi: 10.1111/j.1749-6632.2009.04416.x.

2. Rasch B, Born J. About sleep’s role in memory. Physiol. Rev. 2013;93(2):681–766. doi: 10.1152/physrev.00032.2012.

3. Reutrakul S, Van Cauter E. Sleep influences on obesity, insulin resistance, and risk of type 2 diabetes. Metabolism. 2018;84:56–66. doi: 10.1016/j.metabol.2018.02.010.

4. Chaput J.-P, McHill AW, Cox RC et al. The role of insufficient sleep and circadian misalignment in obesity. Nat. Rev. Endocrinol. 2023;19(2):82–97. doi: 10.1038/s41574-022-00747-7.

5. Benjafield AV, Ayas NT, Eastwood PR et al. Estimation of the global prevalence and burden of obstructive sleep apnoea: a literature-based analysis. Lancet Respir. Med. 2019;7(8):687–698. doi: 10.1016/S2213-2600(19)30198-5.

6. Marin-Oto M, Vicente EE, Marin JM. Long term management of obstructive sleep apnea and its comorbidities. Multidiscip. Respir. Med. 2019;14(1):21. doi: 10.1186/s40248-019-0186-3.

7. Morsy NE, Farrag NS, Zaki NFW et al. Obstructive sleep apnea: personal, societal, public health, and legal implications. Rev. Environ. Health. 2019;34(2):153–169. doi: 10.1007/s10584-019-02764-6.

8. Sata N, Inoshita A, Suda S et al. Clinical, polysomnographic, and cephalometric features of obstructive sleep apnea with AHI over 100. Sleep Breath. 2021;25(3):1379–1387. doi: 10.1007/s11325-020-02241-8.

9. Xia W, Huang Y, Peng B et al. Relationship between obstructive sleep apnoea syndrome and essential hypertension: a dose-response meta-analysis. Sleep Med. 2018;47:11–18. doi: 10.1016/j.sleep.2018.03.016.

10. Zhang X, Fan J, Guo Y et al. Association between obstructive sleep apnoea syndrome and the risk of cardiovascular diseases: an updated systematic review and dose-response meta-analysis. Sleep Med. 2020;71:39–46. doi: 10.1016/j.sleep.2020.03.011.

11. Yu Z, Cheng J.-X, Zhang D, Yi F, Ji Q. Association between obstructive sleep apnea and type 2 diabetes mellitus: A dose-response meta-analysis. Evid. Based. Complement. Alternat. Med. 2021;2021:1337118. doi: 10.1155/2021/1337118.

12. Le Bon O, Staner L, Hoffmann G et al. The first-night effect may last more than one night. J. Psychiatr. Res. 2001;35(3):165–172. doi: 10.1016/s0022-3956(01)00019-x.

13. Markun LC, Sampat A. Clinician-focused overview and developments in polysomnography. Curr. Sleep Med. Rep. 2020;6(4):309–321. doi: 10.1007/s40675-020-00197-5.

14. Kwon S, Kim H, Yeo W.-H. Recent advances in wearable sensors and portable electronics for sleep monitoring. iScience. 2021;24(5):102461. doi: 10.1016/j.isci.2021.102461.

15. Vitazkova D, Kosnacova H, Turonova D et al. Transforming sleep monitoring: Review of wearable and remote devices advancing home polysomnography and their role in predicting neurological disorders. Biosensors (Basel). 2025;15(2):117. doi: 10.3390/bios15020117.

16. Altini M, Kinnunen H. The promise of sleep: A multi-sensor approach for accurate sleep stage detection using the Oura ring. Sensors (Basel). 2021;21(13):4302. doi: 10.3390/s21134302.

17. Fonseca P, Ross M, Cerny A et al. A computationally efficient algorithm for wearable sleep staging in clinical populations. Sci. Rep. 2023;13(1):9182. doi: 10.1038/s41598-023-36444-2.

18. Strumpf Z, Gu W, Tsai C.-W et al. Belun Ring (Belun Sleep System BLS-100): Deep learning-facilitated wearable enables obstructive sleep apnea detection, apnea severity categorization, and sleep stage classification in patients suspected of obstructive sleep apnea. Sleep Health. 2023;9(4):430–440. doi: 10.1016/j.sleh.2023.05.001.

19. Liu P.-K, Ting N, Chiu H.-C et al. Validation of photoplethysmography- and acceleration-based sleep staging in a community sample: comparison with polysomnography and Actiwatch. J. Clin. Sleep Med. 2023;19(10):1797–1810. doi: 10.5664/jcsm.10690.

20. Silva FB, Uribe LFS, Cepeda FX et al. Sleep staging algorithm based on smartwatch sensors for healthy and sleep apnea populations. Sleep Med. 2024;119:535–548. doi: 10.1016/j.sleep.2024.05.033.

21. Constantin L, Horvath CM, Baty F et al. Towards long-term sleep staging via wearable reflective photoplethysmography. Sleep. 2026;49(3):zsaf246. doi: 10.1093/sleep/zsaf246.

22. Imtiaz SA. A systematic review of sensing technologies for wearable sleep staging. Sensors (Basel). 2021;21(5):1562. doi: 10.3390/s21051562.

23. Birrer V, Elgendi M, Lambercy O, Menon C. Evaluating reliability in wearable devices for sleep staging. NPJ Digit. Med. 2024;7(1):74. doi: 10.1038/s41746-024-01016-9.

24. American Academy of Sleep Medicine. The AASM Manual for the Scoring of Sleep and Associated Events: Rules, Terminology and Technical Specifications. Version 3. Darien, IL: American Academy of Sleep Medicine; 2023.

25. Hamaoka T, Murai H, Takata S et al. Different prognosis between severe and very severe obstructive sleep apnea patients; Five year outcomes. J. Cardiol. 2020;76(6):573–579. doi: 10.1016/j.jjcc.2020.06.010.

26. Dai L, Cao W, Luo J, Huang R, Xiao Y. The effectiveness of sleep breathing impairment index in assessing obstructive sleep apnea severity. J. Clin. Sleep Med. 2023;19(2):267–274. doi: 10.5664/jcsm.10302.

27. Korkalainen H, Aakko J, Duce B et al. Deep learning enables sleep staging from photo-plethysmogram for patients with suspected sleep apnea. Sleep. 2020;43(11):zsaa098. doi: 10.1093/sleep/zsaa098.

28. Huttunen R, Leppänen T, Duce B et al. Assessment of obstructive sleep apnea-related sleep fragmentation utilizing deep learning-based sleep staging from photoplethysmography. Sleep. 2021;44(10):zsab142. doi: 10.1093/sleep/zsab142.

29. He K, Zhang X, Ren S, Sun J. Deep residual learning for image recognition. In: 2016 IEEE Conference on Computer Vision and Pattern Recognition (CVPR); Las Vegas, NV, USA; 2016:770–778. doi: 10.1109/cvpr.2016.90.

30. He K, Zhang X, Ren S, Sun J. Identity mappings in deep residual networks. In: Computer Vision – ECCV 2016. Cham: Springer International Publishing; 2016:630–645. doi: 10.1007/978-3-319-46493-0_38.

31. Hochreiter S, Schmidhuber J. Long short-term memory. Neural Comput. 1997;9(8):1735–1780. doi: 10.1162/neco.1997.9.8.1735.

32. Schuster M, Paliwal KK. Bidirectional recurrent neural networks. IEEE Trans. Signal Process. 1997;45(11):2673–2681. doi: 10.1109/78.650093.

33. Lee YJ, Lee JY, Cho JH, Choi JH. Interrater reliability of sleep stage scoring: a meta-analysis. J. Clin. Sleep Med. 2022;18(1):193–202. doi: 10.5664/jcsm.9538.

34. Nikkonen S, Somaskandhan P, Korkalainen H et al. Multicentre sleep-stage scoring agreement in the Sleep Revolution project. J. Sleep Res. 2024;33(1):e13956. doi: 10.1111/jsr.13956.

35. Perslev M, Darkner S, Kempfner L, Nikolic M, Jennum PJ, Igel C. U-Sleep: resilient high-frequency sleep staging. NPJ Digit. Med. 2021;4:72. doi: 10.1038/s41746-021-00440-5.

36. Fine J, Branan KL, Rodriguez AJ et al. Sources of inaccuracy in photoplethysmography for continuous cardiovascular monitoring. Biosensors (Basel). 2021;11(4):126. doi: 10.3390/bios11040126.

37. Johnson DA, Jackson CL, Williams NJ, Alcántara C. Are sleep patterns influenced by race/ethnicity - a marker of relative advantage or disadvantage? Evidence to date. Nat. Sci. Sleep. 2019;11:79–95. doi: 10.2147/NSS.S169312.

38. Sutherland K, Keenan BT, Bittencourt L et al. A global comparison of anatomic risk factors and their relationship to obstructive sleep apnea severity in clinical samples. J. Clin. Sleep Med. 2019;15(4):629–639. doi: 10.5664/jcsm.7730.

39. Quan SF, Howard BV, Iber C et al. The Sleep Heart Health Study: design, rationale, and methods. Sleep. 1997;20(12):1077–1085.

40. Chen X, Wang R, Zee P et al. Racial/ethnic differences in sleep disturbances: The multi-ethnic study of atherosclerosis (MESA). Sleep. 2015;38(6):877–888. doi: 10.5665/sleep.4732.

