## Supplementary Material for "Wearable sleep staging performance declines with sleep apnea severity and is sensitive to training-set composition in very severe sleep apnea"

### A Supplementary Material

#### A.1 Scatter Plots: Associations with Demographic Variables and Sleep Parameters

Scatter plots illustrate associations between sleep staging performance and the following demographic variables and sleep parameters: age, body mass index (BMI), apnea–hypopnea index (AHI), arousal index (ArI), total sleep time (TST), sleep efficiency (SE), wake after sleep onset (WASO), sleep latency (SL), and sleep stage durations (N1, N2, N3, and REM). Performance is quantified using night-level Cohen's kappa for 5-stage sleep staging. We estimated Spearman rank correlation coefficients $\rho$ and report exact two-sided $p$ values together with the correlation coefficients. For reference, we also report Pearson correlation coefficients. These analyses are intended as descriptive exploratory context, and interpretation considered the magnitude and consistency of associations across analyses. Supplementary Table S1 summarizes the correlation analyses for these demographic variables and sleep parameters.

**Table S1:** Correlation between continuous parameters and night-level sleep staging performance.

| Parameter | Dataset | Kappa | | | Macro F1 | | |
|---|---|---|---|---|---|---|---|
| | | Spearman $\rho$ | $p$ | Pearson $r$ | Spearman $\rho$ | $p$ | Pearson $r$ |
| Age | Sleep Lab | -0.239 | $1.6 \times 10^{-5}$ | -0.249 | -0.212 | $2.1 \times 10^{-4}$ | -0.220 |
| | Hospital | -0.273 | $2.3 \times 10^{-5}$ | -0.220 | -0.296 | $3.9 \times 10^{-6}$ | -0.240 |
| BMI | Sleep Lab | -0.232 | $3.0 \times 10^{-5}$ | -0.174 | -0.200 | $3.4 \times 10^{-4}$ | -0.155 |
| | Hospital | -0.091 | 0.164 | -0.105 | -0.096 | 0.144 | -0.110 |
| AHI | Sleep Lab | — | — | — | — | — | — |
| | Hospital | -0.283 | $1.1 \times 10^{-5}$ | -0.360 | -0.267 | $3.4 \times 10^{-5}$ | -0.364 |
| ArI | Sleep Lab | -0.356 | $5.9 \times 10^{-11}$ | -0.458 | -0.319 | $5.8 \times 10^{-9}$ | -0.423 |
| | Hospital | -0.302 | $2.6 \times 10^{-6}$ | -0.345 | -0.290 | $6.6 \times 10^{-6}$ | -0.344 |
| TST | Sleep Lab | 0.158 | $4.6 \times 10^{-3}$ | 0.187 | 0.184 | $9.5 \times 10^{-4}$ | 0.250 |
| | Hospital | 0.095 | 0.148 | 0.094 | 0.156 | 0.017 | 0.157 |
| SE | Sleep Lab | 0.037 | 0.508 | 0.054 | 0.089 | 0.114 | 0.185 |
| | Hospital | 0.100 | 0.126 | 0.083 | 0.181 | $5.6 \times 10^{-3}$ | 0.165 |
| WASO | Sleep Lab | -0.077 | 0.169 | -0.032 | -0.128 | 0.051 | -0.139 |
| | Hospital | -0.130 | 0.020 | -0.078 | -0.175 | $7.2 \times 10^{-3}$ | -0.136 |
| SL | Sleep Lab | 0.076 | 0.175 | 0.118 | -0.045 | 0.496 | 0.069 |
| | Hospital | 0.074 | 0.187 | -0.001 | -0.093 | 0.154 | -0.056 |
| N1 duration | Sleep Lab | -0.331 | $1.4 \times 10^{-8}$ | -0.361 | -0.259 | $3.0 \times 10^{-6}$ | -0.316 |
| | Hospital | -0.330 | $2.4 \times 10^{-7}$ | -0.353 | -0.332 | $2.0 \times 10^{-7}$ | -0.348 |
| N2 duration | Sleep Lab | 0.123 | 0.028 | 0.149 | 0.142 | 0.011 | 0.186 |
| | Hospital | 0.205 | $1.6 \times 10^{-3}$ | 0.208 | 0.228 | $4.2 \times 10^{-4}$ | 0.229 |
| N3 duration | Sleep Lab | 0.228 | $4.1 \times 10^{-5}$ | 0.222 | 0.242 | $1.3 \times 10^{-5}$ | 0.263 |
| | Hospital | 0.368 | $6.7 \times 10^{-9}$ | 0.324 | 0.454 | $2.7 \times 10^{-13}$ | 0.378 |
| REM duration | Sleep Lab | 0.216 | $1.0 \times 10^{-4}$ | 0.265 | 0.215 | $1.1 \times 10^{-4}$ | 0.283 |
| | Hospital | 0.146 | 0.026 | 0.173 | 0.165 | 0.011 | 0.189 |

Performance metrics are night-level Cohen's kappa and macro F1-score for 5-stage sleep staging from cross-validation predictions. Spearman rank correlation $\rho$ with two-sided $p$ values and Pearson correlation $r$ are reported. AHI was evaluated only in the Hospital Dataset.

BMI, body mass index; AHI, apnea–hypopnea index; ArI, arousal index; TST, total sleep time; SE, sleep efficiency; WASO, wake after sleep onset; SL, sleep latency.

Supplementary Figs. S1–S12 summarize these relationships.

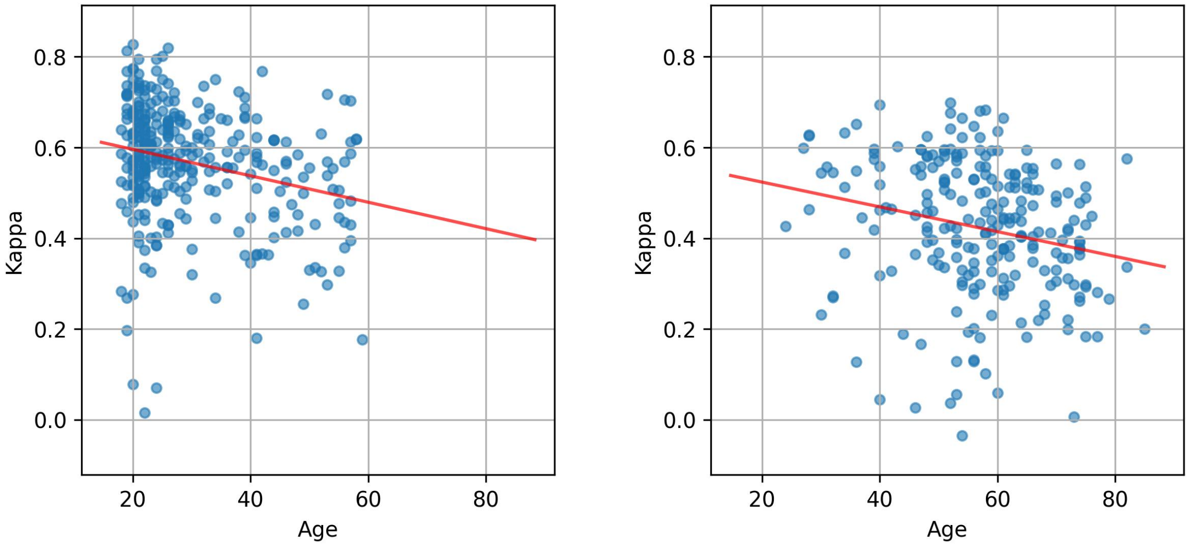


**(a)** Sleep Lab Dataset

**(b)** Hospital Dataset

**Figure S1:** Age (years) vs night-level Cohen's kappa for 5-stage sleep staging. Results are shown for (a) Sleep Lab Dataset and (b) Hospital Dataset. Each point represents one night, and the red line shows the least-squares linear fit.

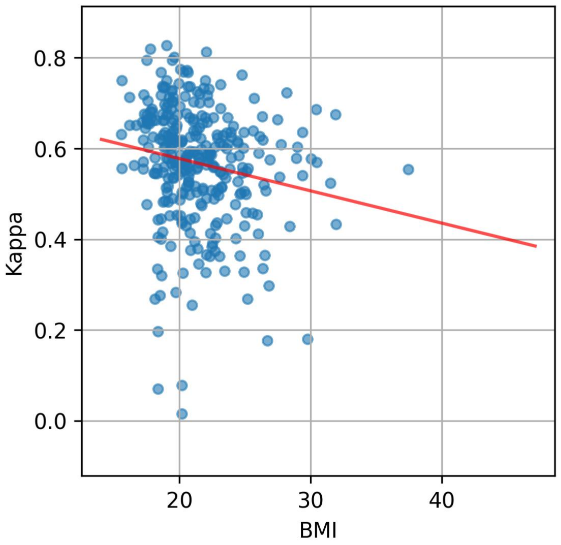


**(a)** Sleep Lab Dataset

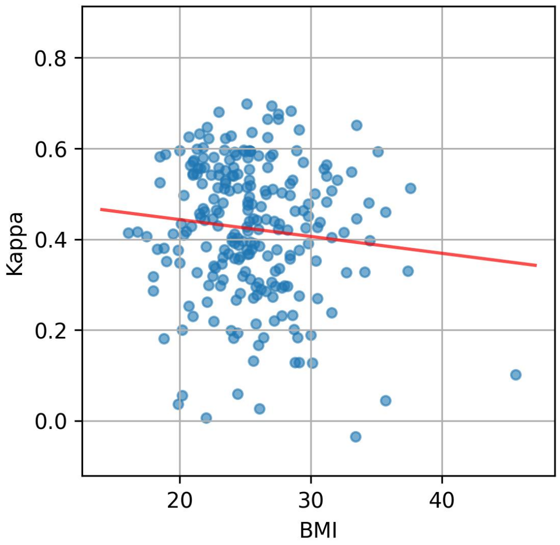


**(b)** Hospital Dataset

**Figure S2:** Body mass index (BMI, kg/m$^2$) vs night-level Cohen's kappa for 5-stage sleep staging. Results are shown for (a) Sleep Lab Dataset and (b) Hospital Dataset. Each point represents one night, and the red line shows the least-squares linear fit.

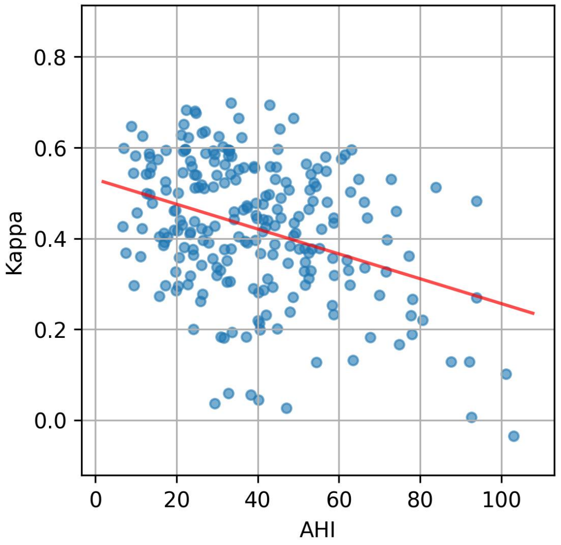


**(a)** Hospital Dataset

**Figure S3:** Apnea–hypopnea index (AHI, events/h) vs night-level Cohen's kappa for 5-stage sleep staging in the Hospital Dataset. Each point represents one night, and the red line shows the least-squares linear fit.

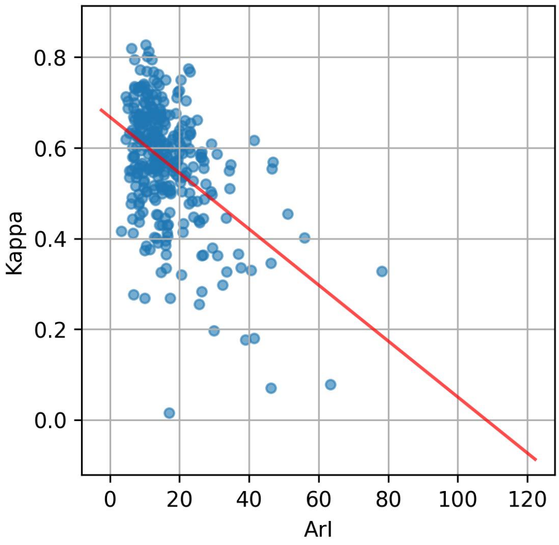


**(a)** Sleep Lab Dataset

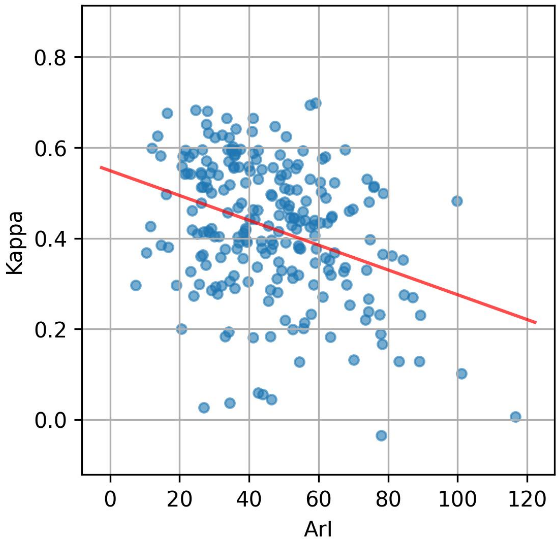


**(b)** Hospital Dataset

**Figure S4:** Arousal index (ArI, events/h) vs night-level Cohen's kappa for 5-stage sleep staging. Results are shown for (a) Sleep Lab Dataset and (b) Hospital Dataset. Each point represents one night, and the red line shows the least-squares linear fit.

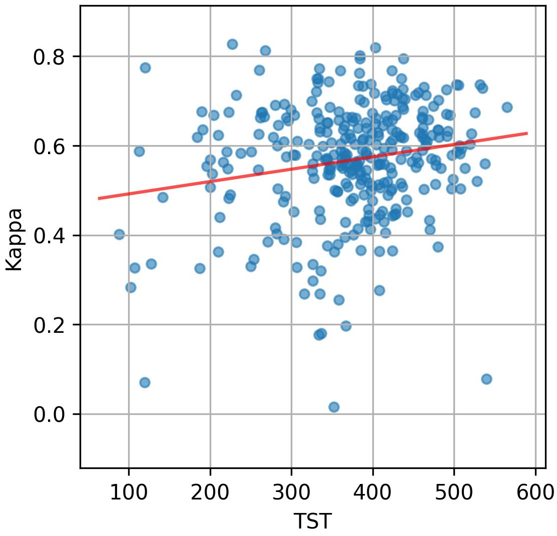


**(a)** Sleep Lab Dataset

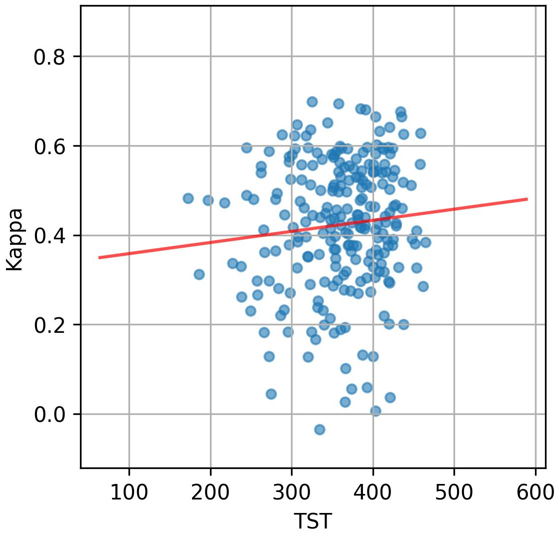


**(b)** Hospital Dataset

**Figure S5:** Total sleep time (TST, min) vs night-level Cohen's kappa for 5-stage sleep staging. Results are shown for (a) Sleep Lab Dataset and (b) Hospital Dataset. Each point represents one night, and the red line shows the least-squares linear fit.

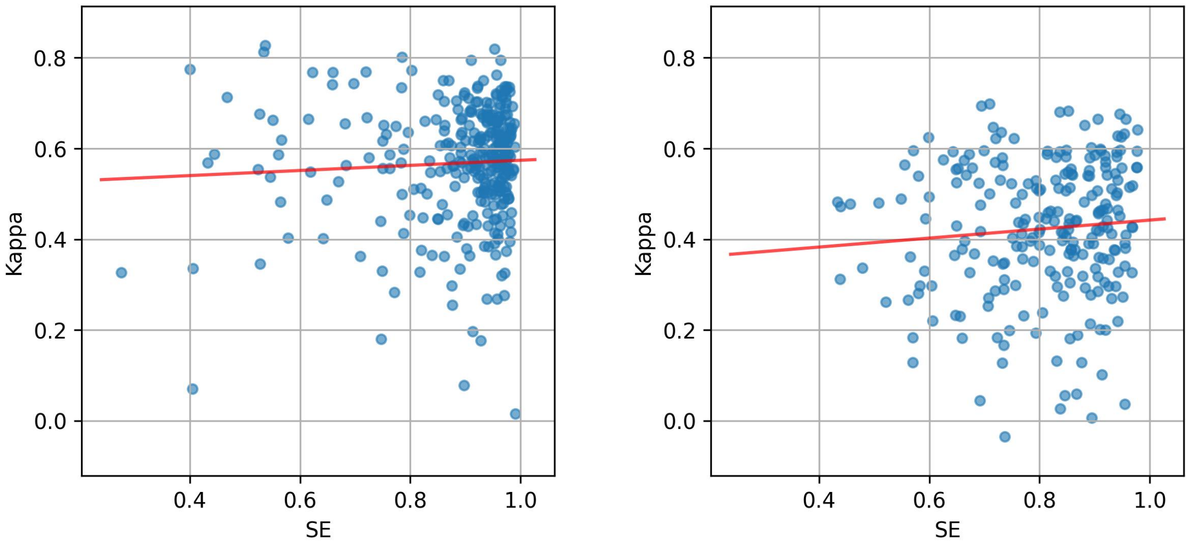


**(a)** Sleep Lab Dataset **(b)** Hospital Dataset

**Figure S6:** Sleep efficiency (SE) vs night-level Cohen's kappa for 5-stage sleep staging. Results are shown for (a) Sleep Lab Dataset and (b) Hospital Dataset. Each point represents one night, and the red line shows the least-squares linear fit.

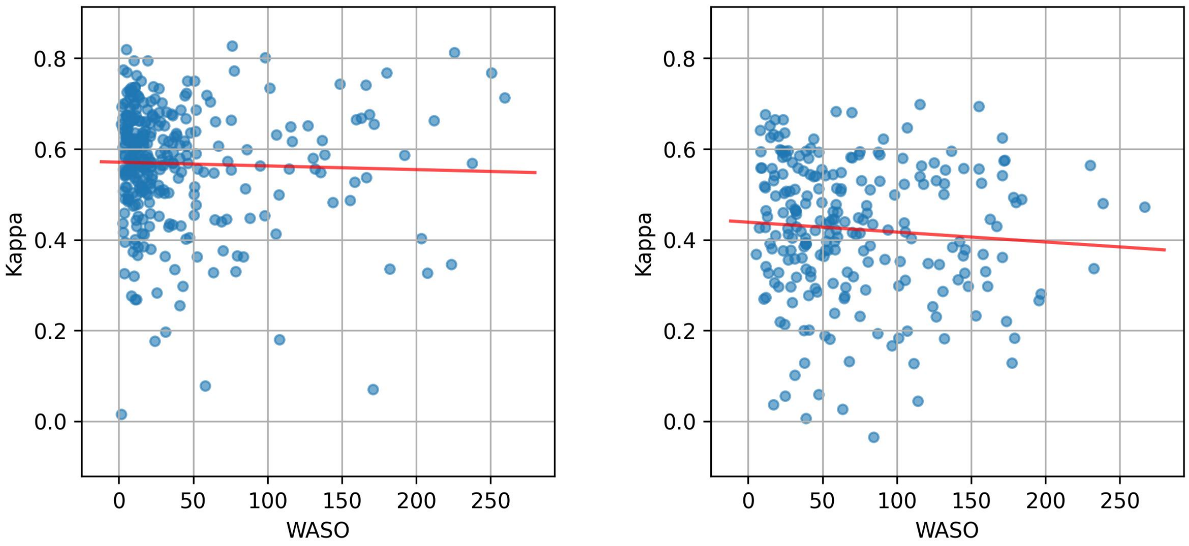


**(a)** Sleep Lab Dataset **(b)** Hospital Dataset

**Figure S7:** Wake after sleep onset (WASO, min) vs night-level Cohen's kappa for 5-stage sleep staging. Results are shown for (a) Sleep Lab Dataset and (b) Hospital Dataset. Each point represents one night, and the red line shows the least-squares linear fit.

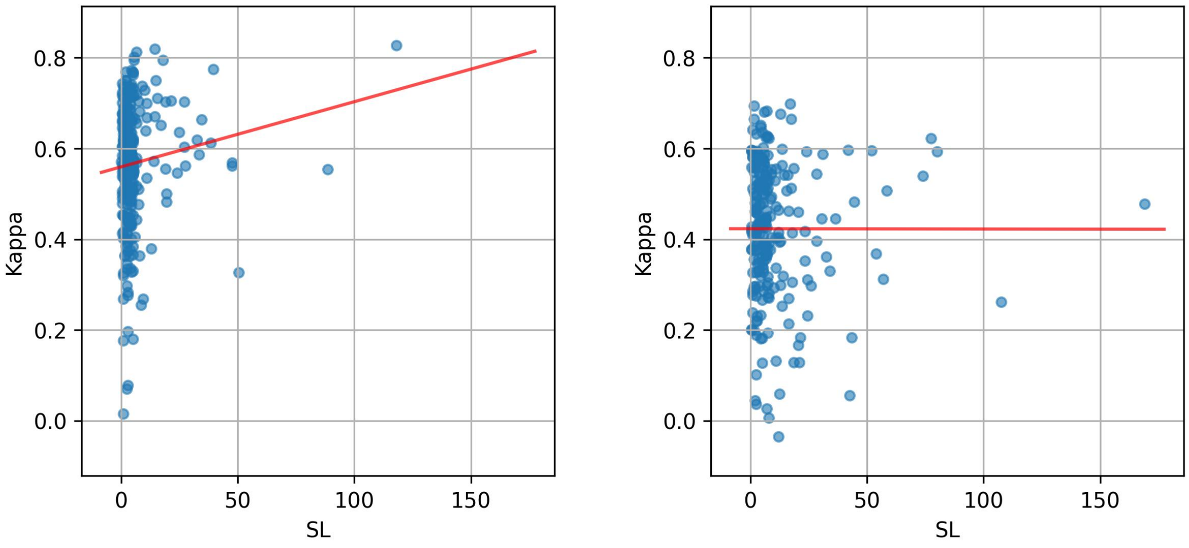


**(a)** Sleep Lab Dataset

**(b)** Hospital Dataset

**Figure S8:** Sleep latency (SL, min) vs night-level Cohen's kappa for 5-stage sleep staging. Results are shown for (a) Sleep Lab Dataset and (b) Hospital Dataset. Each point represents one night, and the red line shows the least-squares linear fit.

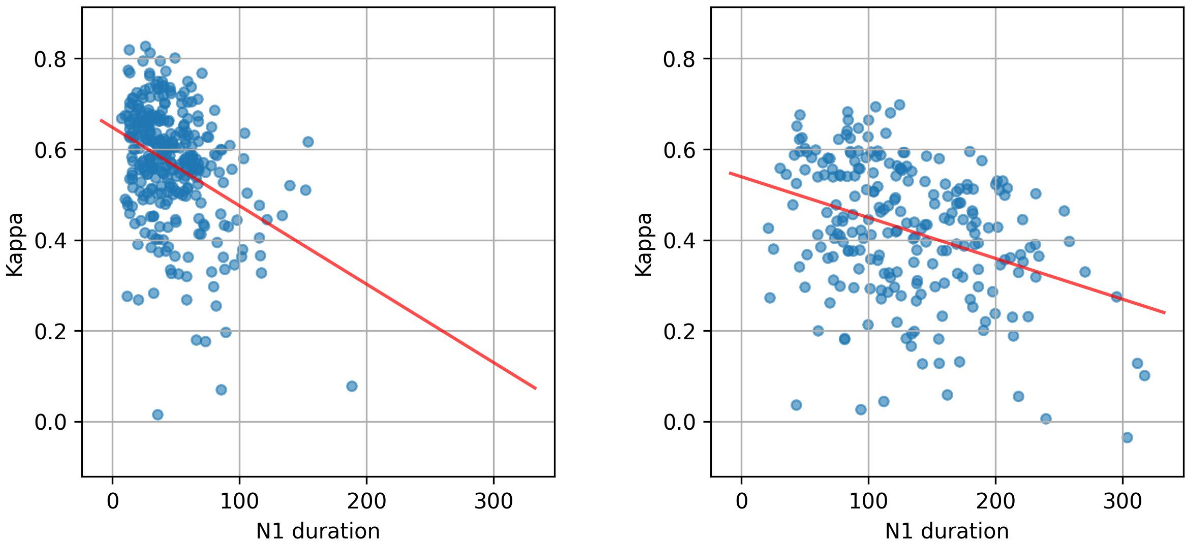


**(a)** Sleep Lab Dataset

**(b)** Hospital Dataset

**Figure S9:** N1 duration (min) vs night-level Cohen's kappa for 5-stage sleep staging. Results are shown for (a) Sleep Lab Dataset and (b) Hospital Dataset. Each point represents one night, and the red line shows the least-squares linear fit.

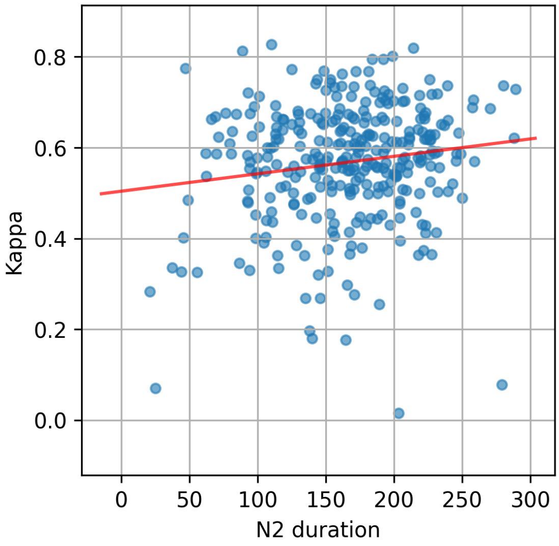


**(a)** Sleep Lab Dataset

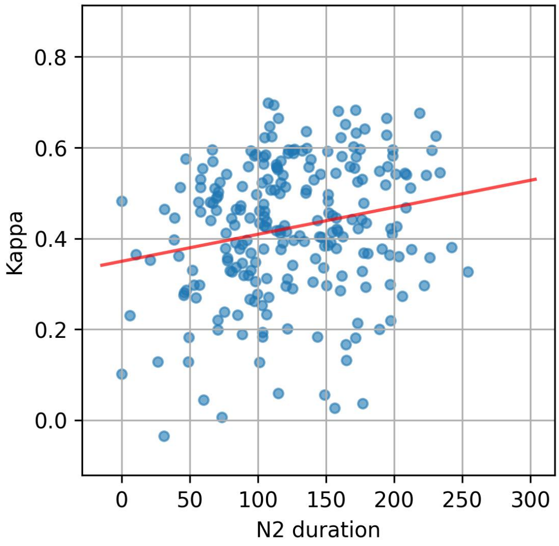


**(b)** Hospital Dataset

**Figure S10:** N2 duration (min) vs night-level Cohen's kappa for 5-stage sleep staging. Results are shown for (a) Sleep Lab Dataset and (b) Hospital Dataset. Each point represents one night, and the red line shows the least-squares linear fit.

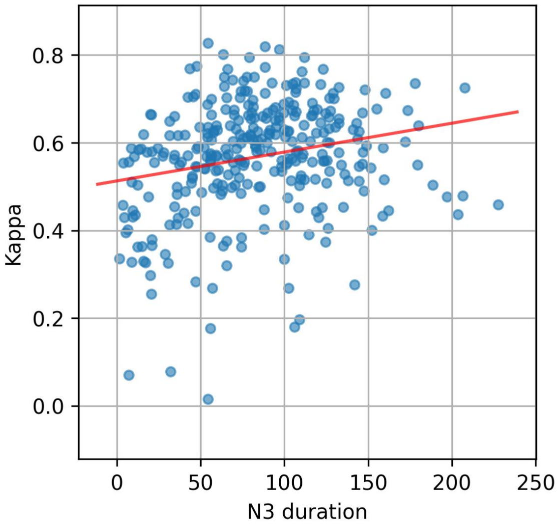


**(a)** Sleep Lab Dataset

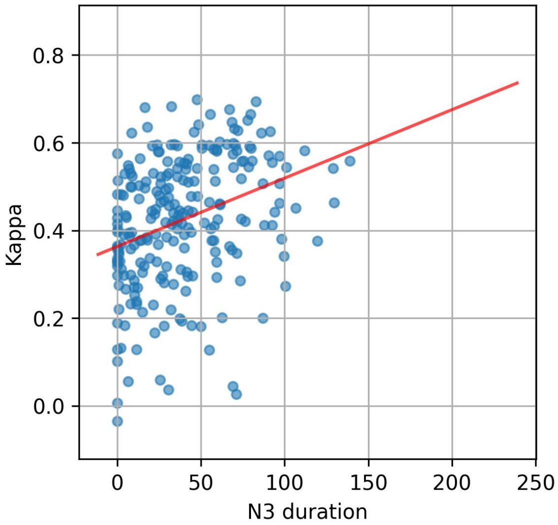


**(b)** Hospital Dataset

**Figure S11:** N3 duration (min) vs night-level Cohen's kappa for 5-stage sleep staging. Results are shown for (a) Sleep Lab Dataset and (b) Hospital Dataset. Each point represents one night, and the red line shows the least-squares linear fit.

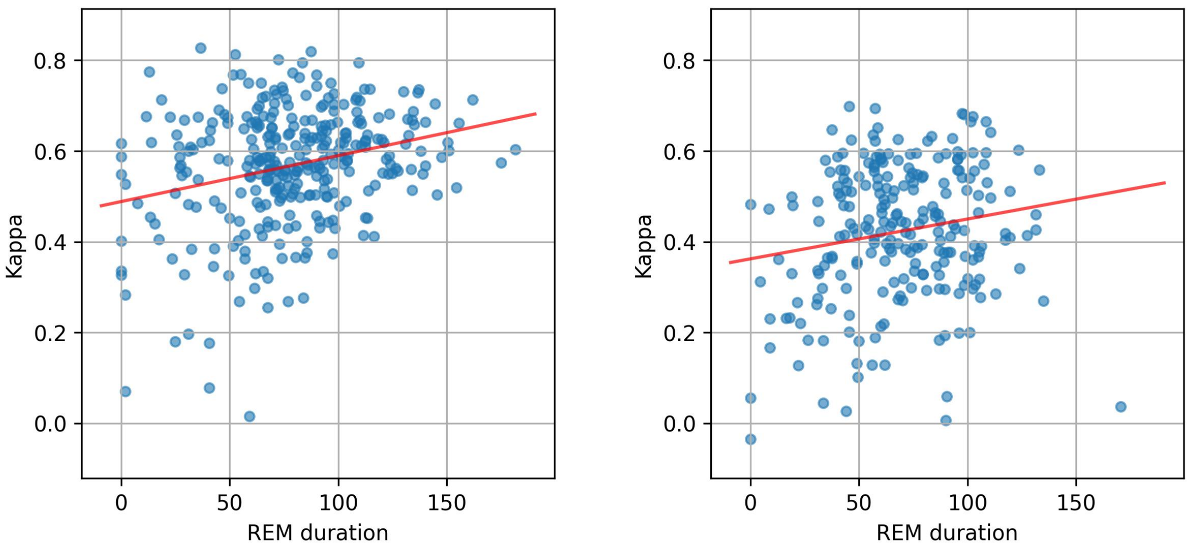


**(a)** Sleep Lab Dataset **(b)** Hospital Dataset

**Figure S12:** REM duration (min) vs night-level Cohen's kappa for 5-stage sleep staging. Results are shown for (a) Sleep Lab Dataset and (b) Hospital Dataset. Each point represents one night, and the red line shows the least-squares linear fit.

#### A.2 Grouped Comparisons for Demographic Variables and Sleep Parameters

This subsection provides grouped comparisons for the demographic variables and sleep parameters other than AHI examined in the main text. The AHI grouped comparison is shown in the main text, whereas the remaining grouped box plots are collected here. Supplementary Figs. S13, S14, and S15 show grouped comparisons for age, sex, and BMI, respectively, and Supplementary Figs. S16–S24 show the corresponding grouped comparisons for ArI, TST, SE, WASO, SL, and sleep-stage durations. These figures are intended to provide descriptive visualization of distribution-level patterns across groups. BMI groups follow the WHO categories [1]. For readability, quartile-based cutpoints shown in the labels were rounded to two significant digits after quartile calculation.

Omnibus Kruskal–Wallis summaries across discretized continuous-variable groups are provided in Supplementary Table S2. To avoid over-interpreting exploratory grouped analyses based on discretized continuous variables, we report omnibus $p$ values and effect sizes ($\epsilon^2$) but do not present exhaustive post hoc pairwise comparisons.

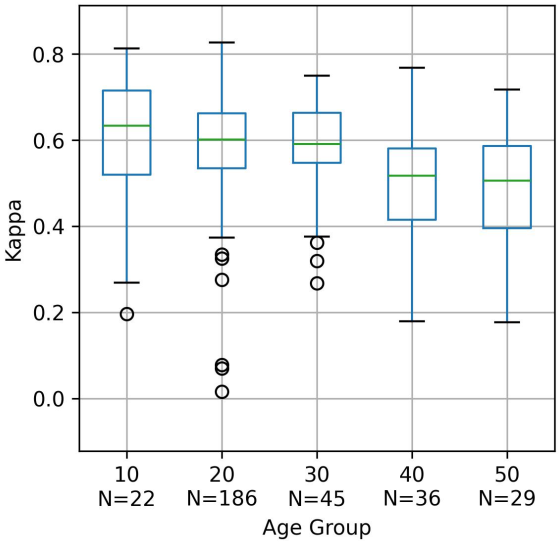


**(a)** Sleep Lab Dataset

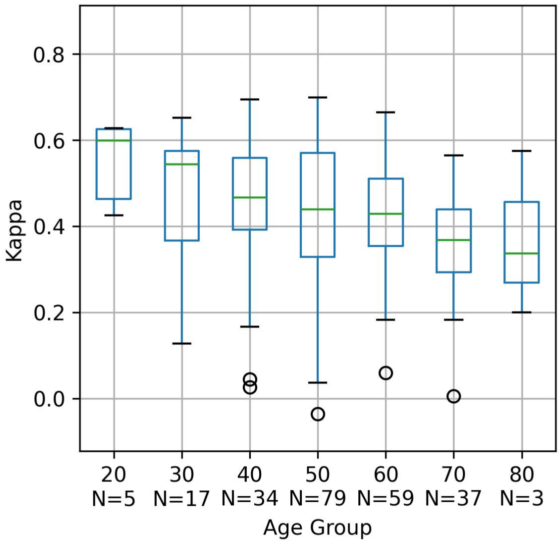


**(b)** Hospital Dataset

**Figure S13:** Sleep staging performance by age group (years, 10-year bins). Results are shown for (a) Sleep Lab Dataset and (b) Hospital Dataset. The y-axis shows night-level Cohen's kappa for 5-stage sleep staging. Group labels indicate the lower bounds of the 10-year bins. Boxes indicate the interquartile range with the median shown as a horizontal line, whiskers extend to 1.5 times the interquartile range, and points indicate outliers.

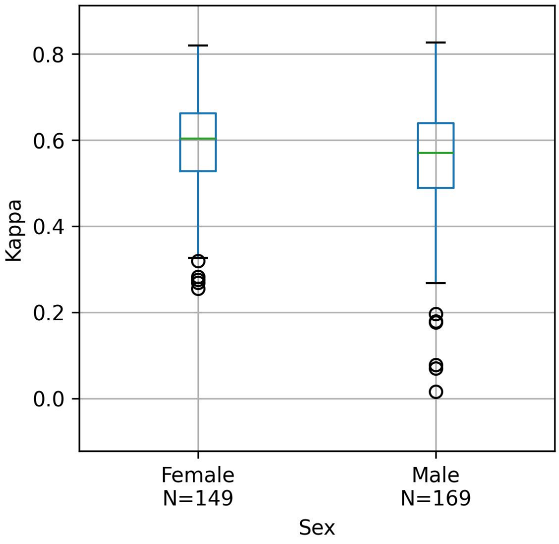


**(a)** Sleep Lab Dataset

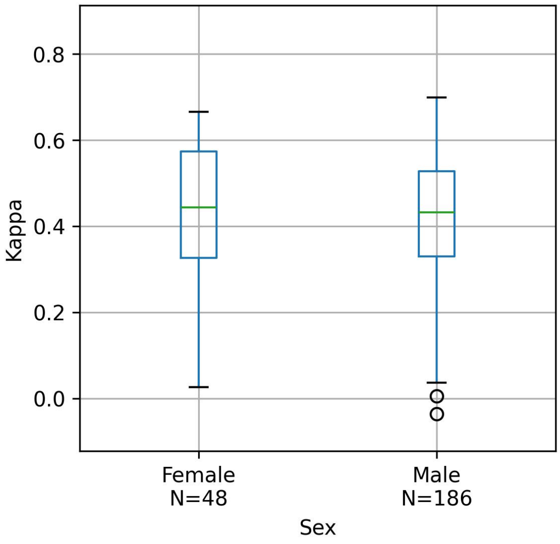


**(b)** Hospital Dataset

**Figure S14:** Sleep staging performance by sex. Results are shown for (a) Sleep Lab Dataset and (b) Hospital Dataset. The y-axis shows night-level Cohen's kappa for 5-stage sleep staging. Boxes indicate the interquartile range with the median shown as a horizontal line, whiskers extend to 1.5 times the interquartile range, and points indicate outliers.

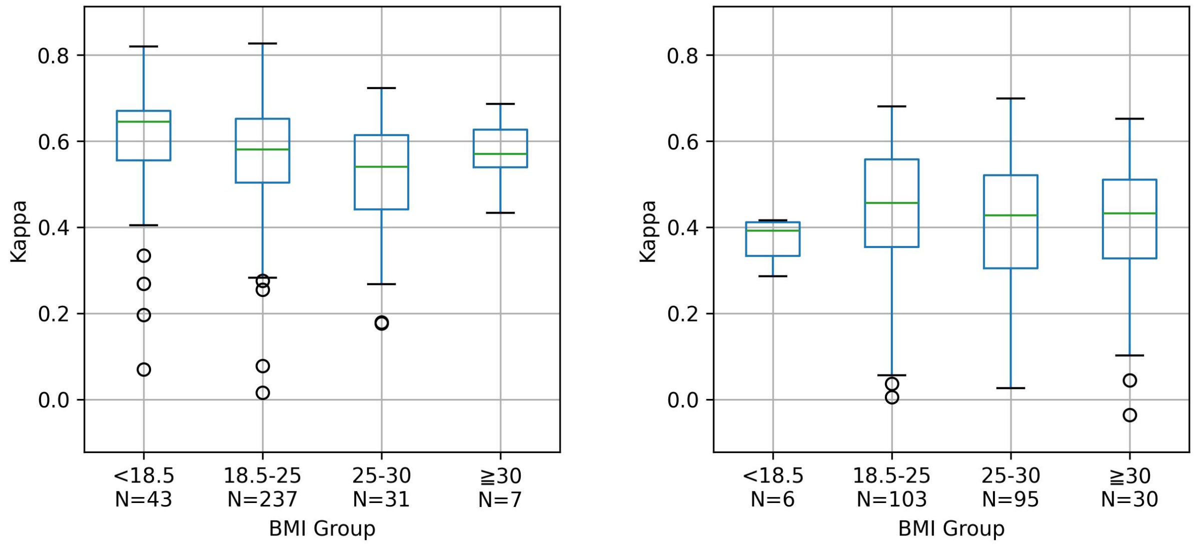


**(a)** Sleep Lab Dataset **(b)** Hospital Dataset

**Figure S15:** Sleep staging performance by body mass index (BMI, kg/m$^2$) group (WHO categories). Results are shown for (a) Sleep Lab Dataset and (b) Hospital Dataset. The y-axis shows night-level Cohen's kappa for 5-stage sleep staging. A group label $x$–$y$ denotes the range $x \leq$ value $< y$. Boxes indicate the interquartile range with the median shown as a horizontal line, whiskers extend to 1.5 times the interquartile range, and points indicate outliers.

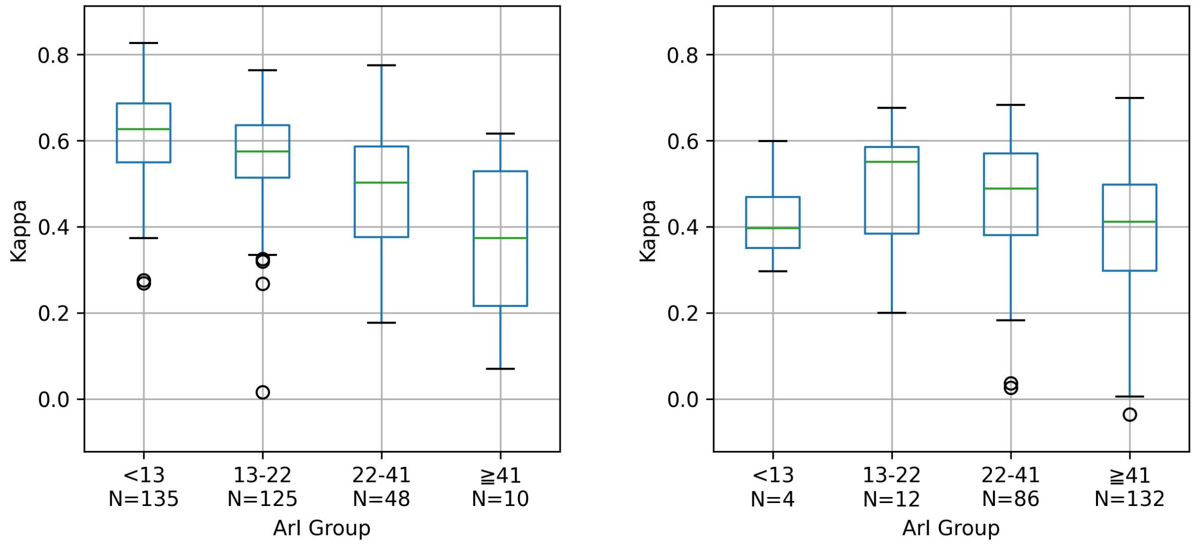


**(a)** Sleep Lab Dataset **(b)** Hospital Dataset

**Figure S16:** Sleep staging performance by arousal index (ArI, events/h) group. Results are shown for (a) Sleep Lab Dataset and (b) Hospital Dataset. The y-axis shows night-level Cohen's kappa for 5-stage sleep staging. Groups correspond to quartiles computed from the pooled Sleep Lab Dataset and Hospital Dataset, and the same cutpoints were applied to both panels. A group label $x$–$y$ denotes the range $x \leq$ value $< y$. Boxes indicate the interquartile range with the median shown as a horizontal line, whiskers extend to 1.5 times the interquartile range, and points indicate outliers.

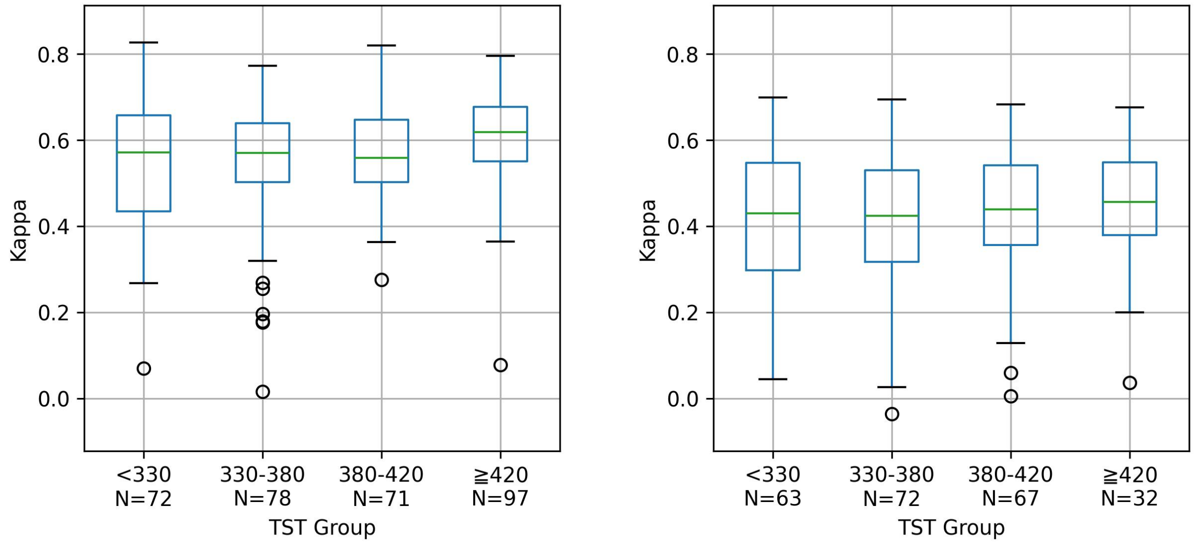


**(a)** Sleep Lab Dataset **(b)** Hospital Dataset

**Figure S17:** Sleep staging performance by total sleep time (TST, min) group. Results are shown for (a) Sleep Lab Dataset and (b) Hospital Dataset. The y-axis shows night-level Cohen's kappa for 5-stage sleep staging. Groups correspond to quartiles computed from the pooled Sleep Lab Dataset and Hospital Dataset, and the same cutpoints were applied to both panels. A group label $x$–$y$ denotes the range $x \leq$ value $< y$. Boxes indicate the interquartile range with the median shown as a horizontal line, whiskers extend to 1.5 times the interquartile range, and points indicate outliers.

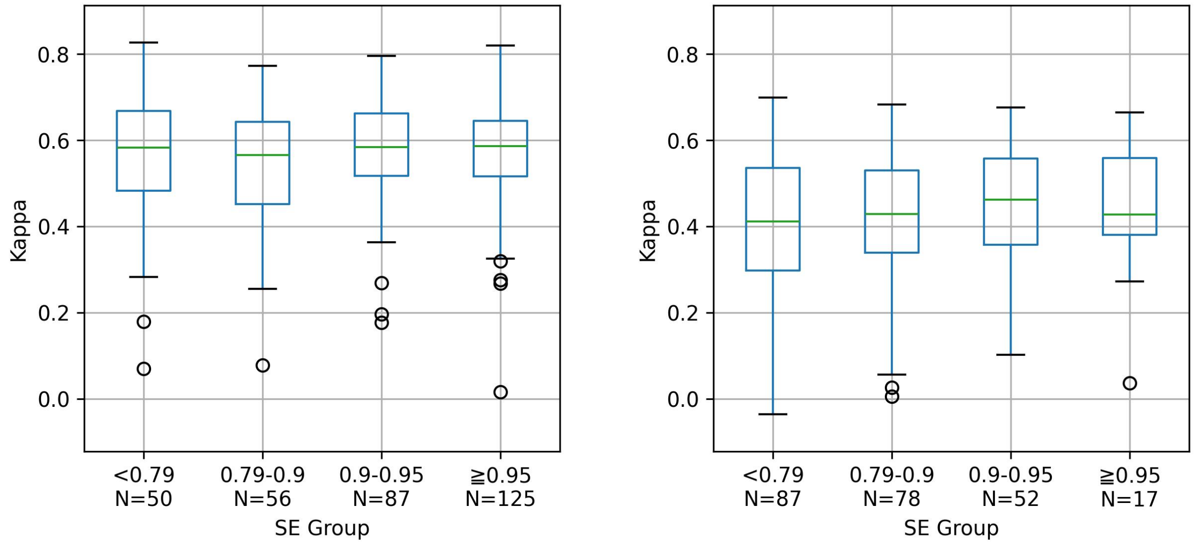


**(a)** Sleep Lab Dataset **(b)** Hospital Dataset

**Figure S18:** Sleep staging performance by sleep efficiency (SE) group. Results are shown for (a) Sleep Lab Dataset and (b) Hospital Dataset. The y-axis shows night-level Cohen's kappa for 5-stage sleep staging. Groups correspond to quartiles computed from the pooled Sleep Lab Dataset and Hospital Dataset, and the same cutpoints were applied to both panels. A group label $x$–$y$ denotes the range $x \leq$ value $< y$. Boxes indicate the interquartile range with the median shown as a horizontal line, whiskers extend to 1.5 times the interquartile range, and points indicate outliers.

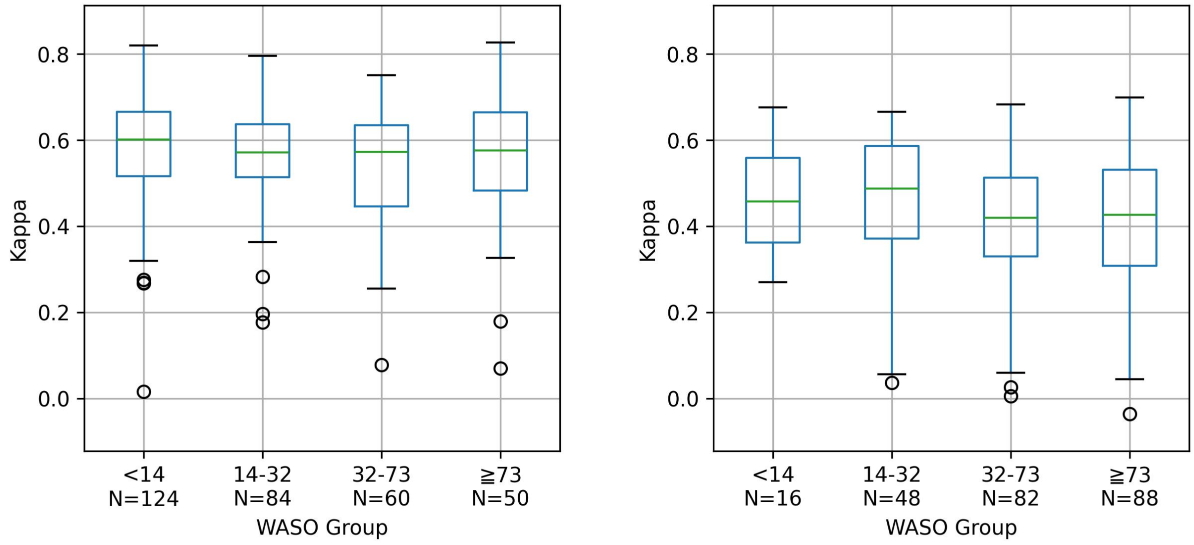


**(a)** Sleep Lab Dataset

**(b)** Hospital Dataset

**Figure S19:** Sleep staging performance by wake after sleep onset (WASO, min) group. Results are shown for (a) Sleep Lab Dataset and (b) Hospital Dataset. The y-axis shows night-level Cohen's kappa for 5-stage sleep staging. Groups correspond to quartiles computed from the pooled Sleep Lab Dataset and Hospital Dataset, and the same cutpoints were applied to both panels. A group label $x$–$y$ denotes the range $x \leq$ value $< y$. Boxes indicate the interquartile range with the median shown as a horizontal line, whiskers extend to 1.5 times the interquartile range, and points indicate outliers.

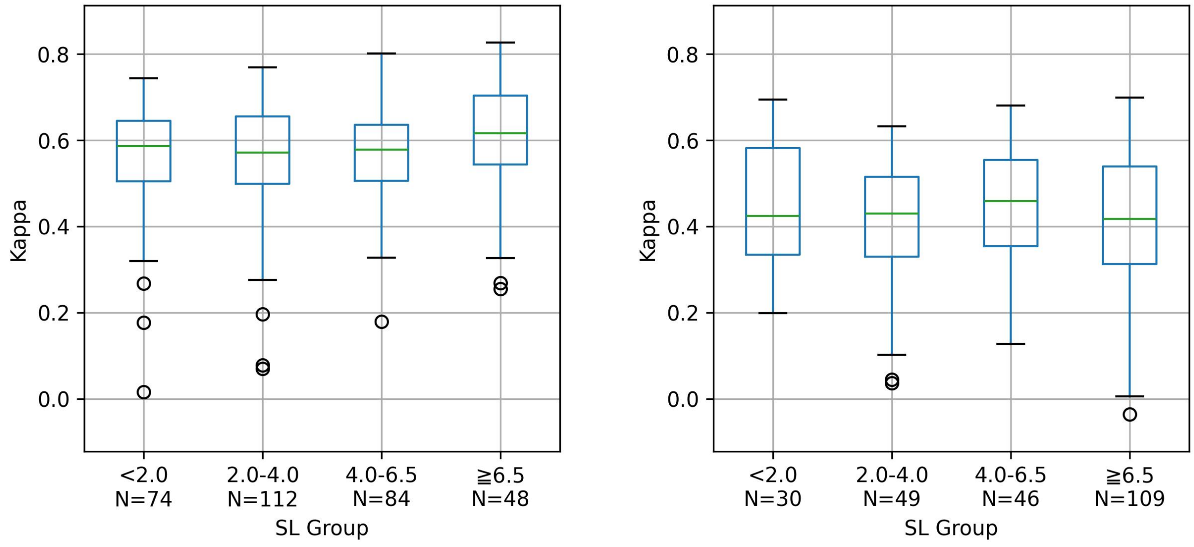


**(a)** Sleep Lab Dataset

**(b)** Hospital Dataset

**Figure S20:** Sleep staging performance by sleep latency (SL, min) group. Results are shown for (a) Sleep Lab Dataset and (b) Hospital Dataset. The y-axis shows night-level Cohen's kappa for 5-stage sleep staging. Groups correspond to quartiles computed from the pooled Sleep Lab Dataset and Hospital Dataset, and the same cutpoints were applied to both panels. A group label $x$–$y$ denotes the range $x \leq$ value $< y$. Boxes indicate the interquartile range with the median shown as a horizontal line, whiskers extend to 1.5 times the interquartile range, and points indicate outliers.

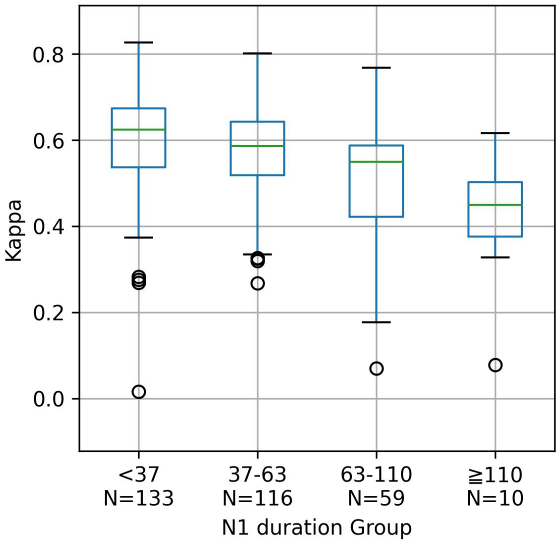


**(a)** Sleep Lab Dataset

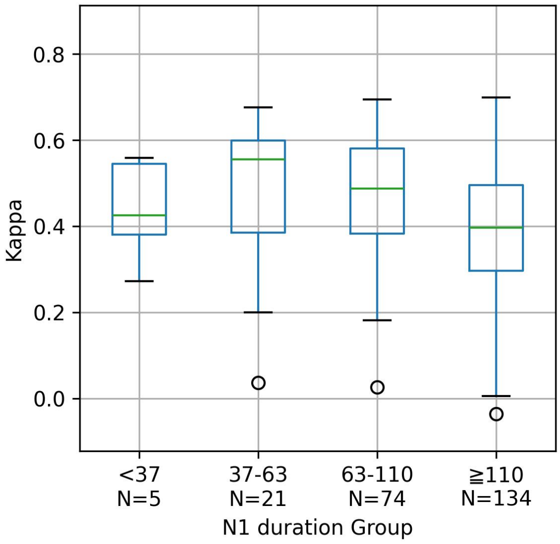


**(b)** Hospital Dataset

**Figure S21:** Sleep staging performance by N1 duration (min) group. Results are shown for (a) Sleep Lab Dataset and (b) Hospital Dataset. The y-axis shows night-level Cohen's kappa for 5-stage sleep staging. Groups correspond to quartiles computed from the pooled Sleep Lab Dataset and Hospital Dataset, and the same cutpoints were applied to both panels. A group label $x$–$y$ denotes the range $x \leq$ value $< y$. Boxes indicate the interquartile range with the median shown as a horizontal line, whiskers extend to 1.5 times the interquartile range, and points indicate outliers.

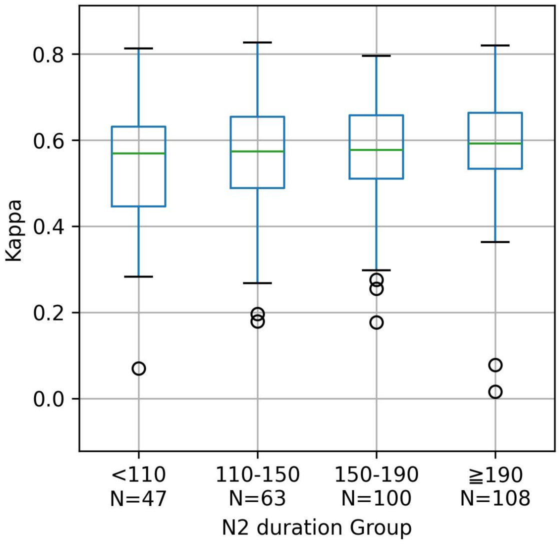


**(a)** Sleep Lab Dataset

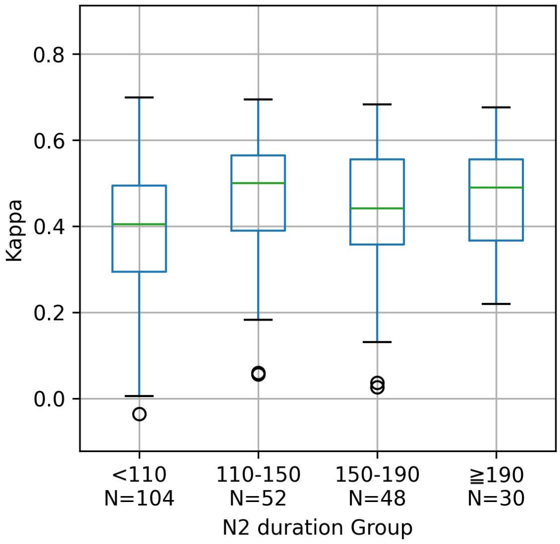


**(b)** Hospital Dataset

**Figure S22:** Sleep staging performance by N2 duration (min) group. Results are shown for (a) Sleep Lab Dataset and (b) Hospital Dataset. The y-axis shows night-level Cohen's kappa for 5-stage sleep staging. Groups correspond to quartiles computed from the pooled Sleep Lab Dataset and Hospital Dataset, and the same cutpoints were applied to both panels. A group label $x$–$y$ denotes the range $x \leq$ value $< y$. Boxes indicate the interquartile range with the median shown as a horizontal line, whiskers extend to 1.5 times the interquartile range, and points indicate outliers.

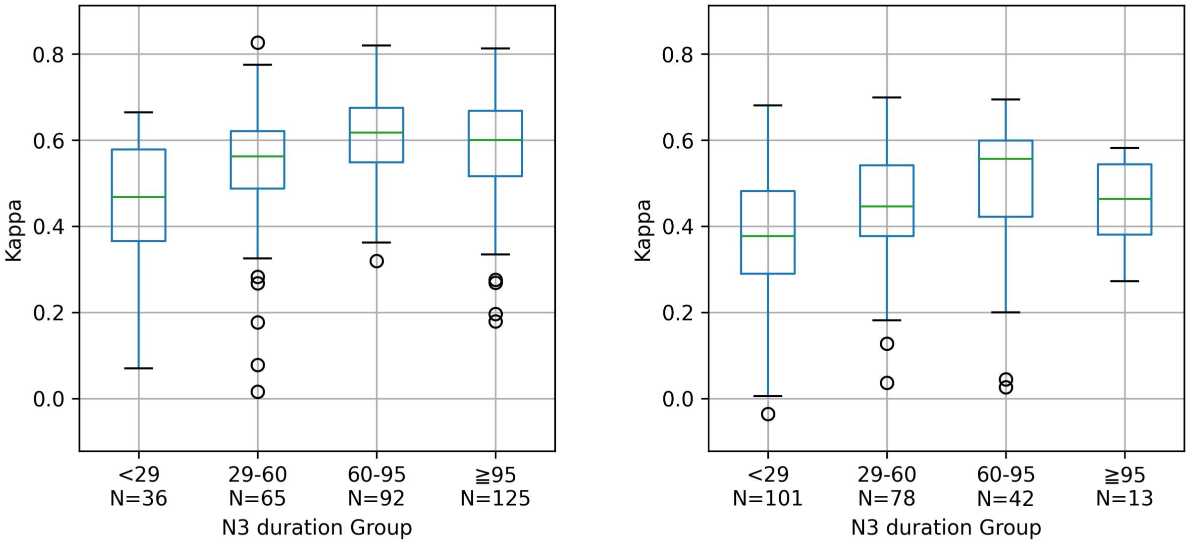


**(a)** Sleep Lab Dataset **(b)** Hospital Dataset

**Figure S23:** Sleep staging performance by N3 duration (min) group. Results are shown for (a) Sleep Lab Dataset and (b) Hospital Dataset. The y-axis shows night-level Cohen's kappa for 5-stage sleep staging. Groups correspond to quartiles computed from the pooled Sleep Lab Dataset and Hospital Dataset, and the same cutpoints were applied to both panels. A group label $x$–$y$ denotes the range $x \leq$ value $< y$. Boxes indicate the interquartile range with the median shown as a horizontal line, whiskers extend to 1.5 times the interquartile range, and points indicate outliers.

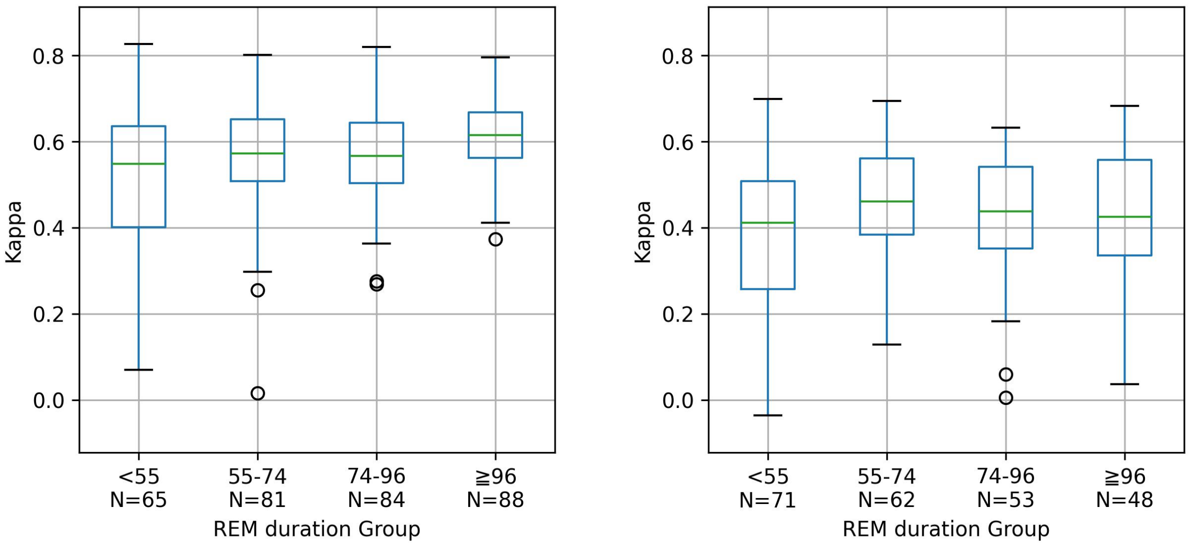


**(a)** Sleep Lab Dataset **(b)** Hospital Dataset

**Figure S24:** Sleep staging performance by REM duration (min) group. Results are shown for (a) Sleep Lab Dataset and (b) Hospital Dataset. The y-axis shows night-level Cohen's kappa for 5-stage sleep staging. Groups correspond to quartiles computed from the pooled Sleep Lab Dataset and Hospital Dataset, and the same cutpoints were applied to both panels. A group label $x$–$y$ denotes the range $x \leq$ value $< y$. Boxes indicate the interquartile range with the median shown as a horizontal line, whiskers extend to 1.5 times the interquartile range, and points indicate outliers.

**Table S2:** Comparison of night-level sleep staging performance across discretized continuous-parameter groups.

| Parameter | Dataset | Kappa | | Macro F1 | |
|---|---|---|---|---|---|
| | | Kruskal-Wallis $p$ | $\epsilon^2$ | Kruskal-Wallis $p$ | $\epsilon^2$ |
| Age | Sleep Lab | $6.5 \times 10^{-6}$ | 0.093 | $1.7 \times 10^{-4}$ | 0.071 |
| | Hospital | $4.5 \times 10^{-3}$ | 0.081 | $9.7 \times 10^{-4}$ | 0.097 |
| Sex | Sleep Lab | 0.052 | 0.012 | 0.100 | 0.009 |
| | Hospital | 0.349 | 0.004 | 0.277 | 0.005 |
| BMI | Sleep Lab | 0.028 | 0.029 | 0.277 | 0.012 |
| | Hospital | 0.280 | 0.017 | 0.251 | 0.018 |
| AHI | Sleep Lab | — | — | — | — |
| | Hospital | $5.2 \times 10^{-4}$ | 0.076 | $8.0 \times 10^{-4}$ | 0.072 |
| ArI | Sleep Lab | $3.2 \times 10^{-10}$ | 0.149 | $2.0 \times 10^{-7}$ | 0.107 |
| | Hospital | $7.8 \times 10^{-4}$ | 0.072 | $1.4 \times 10^{-3}$ | 0.067 |
| TST | Sleep Lab | $9.6 \times 10^{-3}$ | 0.036 | $2.7 \times 10^{-3}$ | 0.045 |
| | Hospital | 0.582 | 0.008 | 0.171 | 0.022 |
| SE | Sleep Lab | 0.680 | 0.005 | 0.340 | 0.011 |
| | Hospital | 0.391 | 0.013 | 0.041 | 0.036 |
| WASO | Sleep Lab | 0.435 | 0.009 | 0.134 | 0.018 |
| | Hospital | 0.162 | 0.022 | 0.084 | 0.029 |
| SL | Sleep Lab | 0.172 | 0.016 | 0.095 | 0.020 |
| | Hospital | 0.474 | 0.011 | 0.246 | 0.018 |
| N1 duration | Sleep Lab | $1.3 \times 10^{-7}$ | 0.110 | $6.4 \times 10^{-6}$ | 0.085 |
| | Hospital | $1.8 \times 10^{-5}$ | 0.106 | $3.8 \times 10^{-5}$ | 0.099 |
| N2 duration | Sleep Lab | 0.188 | 0.015 | 0.052 | 0.024 |
| | Hospital | $9.8 \times 10^{-3}$ | 0.049 | 0.014 | 0.046 |
| N3 duration | Sleep Lab | $1.4 \times 10^{-7}$ | 0.110 | $2.2 \times 10^{-7}$ | 0.107 |
| | Hospital | $2.4 \times 10^{-6}$ | 0.124 | $2.4 \times 10^{-9}$ | 0.185 |
| REM duration | Sleep Lab | $1.9 \times 10^{-3}$ | 0.047 | $1.1 \times 10^{-3}$ | 0.050 |
| | Hospital | 0.045 | 0.035 | 0.013 | 0.046 |

Performance metrics are night-level Cohen's kappa and macro F1-score for 5-stage sleep staging from cross-validation predictions. Kruskal–Wallis $p$ values and effect sizes $\epsilon^2$ are reported.
BMI, body mass index; AHI, apnea–hypopnea index; ArI, arousal index; TST, total sleep time; SE, sleep efficiency; WASO, wake after sleep onset; SL, sleep latency.

### References

1. World Health Organization. Obesity: preventing and managing the global epidemic. Report of a WHO consultation. World Health Organ. Tech. Rep. Ser. 2000;894:i–xii, 1–253.
